# Discriminatory cytokine profiles predict muscle function, fatigue and cognitive function in patients with Myalgic Encephalomyelitis/Chronic Fatigue Syndrome (ME/CFS)

**DOI:** 10.1101/2020.08.17.20164715

**Authors:** A Gusnanto, KE Earl, GK Sakellariou, DJ Owens, A Lightfoot, S Fawcett, E Owen, CA Staunton, T Shu, FC Croden, M Fenech, M Sinclair, L Ratcliffe, KA Whysall, R Haynes, NM Wells, MJ Jackson, GL Close, C Lawton, MBJ Beadsworth, L Dye, A McArdle

## Abstract

Myalgic Encephalomyelitis (ME) /Chronic Fatigue Syndrome (CFS) is a severely debilitating and complex illness of uncertain aetiology, affecting the lives of millions and characterised by prolonged fatigue. The initiating factors and mechanisms leading to chronic debilitating muscle fatigue in ME/CFS are unknown and are complicated by the time required for diagnosis. Both mitochondrial dysfunction and inflammation have been proposed to be central to the pathogenesis of ME/CFS. This original and extensive study demonstrated that although there was little dysfunction evident in the muscle mitochondria of patients with ME/CFS, particular blood plasma and skeletal muscle cytokines, when adjusted for age, gender and cytokine interactions could predict both diagnosis and a number of measures common to patients with ME/CFS. These included MVC and perceived fatigue as well as cognitive indices such as pattern and verbal reaction times. We employed advanced multivariate analyses to cytokine profiles that leverages covariation and intrinsic redundancy to identify patterns of immune signaling that can be evaluated for their predictions of disease phenotype. The current study identified discriminatory cytokine profiles that can be sufficiently used to distinguish HCs from patients with ME/CFS and provides compelling evidence that a limited number of cytokines are associated with diagnosis and fatigue. Moreover, this study demonstrates significant potential of using multiplex cytokine profiles and bioinformatics as diagnostic tools for ME/CFS, potentiating the possibility of not only diagnosis, but also being able to individually personalise therapies.

**Significance:** Myalgic Encephalomyelitis/Chronic Fatigue Syndrome (ME/CFS) is a complex, chronic, debilitating and potentially life-changing medical condition affecting children and adults of all ages, races and socio-economic groupings. Clinical presentation includes fluctuating fatigue of varying severity, with other symptoms, including myalgia, arthralgia, post-exertional fatigue, unrefreshed sleep, headache, upper respiratory tract symptoms, and cognitive impairment. With no biomarkers, or diagnostic tests, aetiology, epidemiology and pathophysiology remain unclear. This extensive study employed advanced multivariate analyses that leveraged covariation and intrinsic redundancy and identified discriminatory cytokine profiles that can be used to distinguish Healthy Controls (HCs) from patients with ME/CFS and a limited number of cytokines were associated with physical and cognitive fatigue. These findings are relevant to the potential of increasing numbers of patients developing chronic fatigue following Coronavirus disease 2019.

## Introduction

Myalgic Encephalomyelitis/Chronic Fatigue Syndrome (ME/CFS) is a complex, chronic, debilitating and potentially life-changing medical condition (1). It affects children and adults of all ages, races and socio-economic groupings. Clinical presentation includes fluctuating fatigue of varying severity, with a range of other symptoms, including myalgia, arthralgia, post-exertional fatigue, unrefreshed sleep, headache, upper respiratory tract symptoms, and cognitive impairment (2). With no current biomarkers, or diagnostic tests, and sparse funding of research (3), aetiology, epidemiology and pathophysiology remain to be determined. National Institute for Health and Care Excellence (NICE) guidelines ‘confirm the lack of epidemiological data, for the UK and suggest a population prevalence of at least 0.2–0.4%.’(1), others estimate prevalence between 0.2 and 1% (4-6).

Both initiating and precipitating factors and mechanisms leading to the chronic debilitating muscle fatigue in patients with ME/CFS are unknown. There are no established biomarkers in ME/CFS, but there has been a long-standing interest in the role of the immune system in ME/CFS pathogenesis and the role that cytokines might play in disease development (7). Onset of ME/CFS often follows an acute infectious episode, particularly of infectious mononucleosis and the persistence of ‘flu-like’ symptoms has led to the suggestion that the illness is due to an inflammatory disorder (8-12). There are at least 20 ‘consensus’ clinical or research definitions for ME/CFS (13). There is heterogeneity amongst them, however, chronicity of symptoms is key to making the diagnosis. NICE guidelines suggest greater than four months, whilst the Oxford-1991 and CDC-1994-Fukuda, the most frequently used definitions in research require greater than six months. Thus, the time required for diagnosis further complicates the identification of factors responsible for initiation of ME/CFS. This is particularly relevant for skeletal muscle since several months of reduced activity causes a reduction in muscle mass and function that may compound or mask underlying aetiological factors.

The mechanisms by which fatigue develops remain elusive (14, 15). Despite the issues with diagnosis of ME/CFS, it has been proposed that there is a core condition to ME/CFS that involves mitochondrial abnormalities and/or inflammation. A failure of appropriate energy supply to muscles is a fundamental and direct cause of fatigue thus it is not surprising that defective mitochondria have been implicated in conditions where fatigue is prevalent such as ageing, cardiovascular disease, neurodegenerative disorders (16) as well as ME/CFS (17). In addition to being the major source of energy for sustained muscle contractions, mitochondria generate reactive oxygen species (ROS) during normal respiration. Skeletal muscle fibres respond to contractile activity by increasing intracellular generation of superoxide and nitric oxide with the formation of secondary ROS and reactive nitrogen species (18-20). ROS scavengers have been reported to delay fatigue (21), supporting the hypothesis that physiological generation of ROS contributes to fatigue. A mouse model with aberrant mitochondrial ROS generation, the superoxide dismutase 2 knockout (SOD2KO) mouse, shows severe disturbances in exercise activity associated with increased oxidative damage and reduced ATP content in their muscle tissue (22, 23), suggesting that aberrant superoxide radical generated by mitochondria play a significant role in exercise tolerance. Mitochondrial dysfunction in the central nervous system has been implicated in a wide range of mental or neurological conditions, including chronic psychological stress and fatigue, cognitive deficits, anxiety and depression (24). Rodent studies support this, for example, mouse studies have demonstrated that mitochondrial ROS contribute to age-related cerebro-microvascular dysfunction and that preservation of mitochondria with a mitochondrial targeted antioxidant preserved memory (25). A study in rats has demonstrated chemotherapy-induced mitochondrial dysfunction was associated with cognitive impairments as well as neuropathy. Moreover, antioxidant treatment prevented the drug-induced mitochondrial changes and partially reversed the cognitive impairments (26).

Whether mitochondria are abnormal in patients with ME/CFS remains the subject of considerable debate. Behan et al. (27) demonstrated substantial mitochondrial structural abnormalities in muscles of patients, including branching and fusion of crystae, swelling, vacuolation, myelin figures and secondary lysosomes. Myhill et al (28) found a significant correlation between the degree of mitochondrial dysfunction in blood cells and the severity of illness. In contrast, Plioplys and Plioplys (29) and Barnes et al (30) failed to demonstrate any specific abnormalities in skeletal muscles of patients with ME/CFS. Contrasting findings are likely to be due to a number of factors and particularly the limited methods so far applied to analyse mitochondrial function in patients with ME/CFS. Few studies have used sensitive techniques to determine mitochondrial function directly in muscle fibres where results would not be influenced by the exercise history of the patients or their ability to sustain an experimental exercise protocol.

Together with mitochondrial dysfunction, chronic inflammation is a hallmark of a number of conditions, particularly where patients complain of fatigue, including ME/CFS (31-34). Studies have shown that plasma levels of cytokines are chronically modified in patients with CFS (35-37) and that limited numbers of cytokines are associated with severity of disease and fatigue duration (38). Chronic NFκ-B activation, particularly in blood cells, is proposed as a key event in ME/CFS, leading to a pro-inflammatory immune response (39). The mechanism(s) by which activation occurs are unclear but symptoms of ME/CFS may be due to this chronic inflammatory response (39).

The impact of chronic and systemic inflammation on skeletal muscle function is profound and includes the development of atrophy, weakness and fatigue (40). Similarly, changes in plasma levels of cytokines are associated with modified learning and memory (41-43). It has been proposed that mitochondrial dysfunction and chronic inflammation may be linked such that mitochondrial dysregulation or damage results in systemic inflammation by the cellular release of mitochondrial components (16). Conversely, we and others have demonstrated that changes in extracellular cytokines, such as TNF-α results in increased mitochondrial ROS generation in muscle fibres with subsequent release of additional cytokines by muscle cells (44).

The purpose of the current study was to comprehensively assess muscle function and plasma immune profiles in patients with ME/CFS and to use newly developed techniques to permit examination of skeletal muscle mitochondrial function *in situ*. The study also investigated the potential contribution of muscle-derived cytokines to the plasma immune profile and explored differences in cognitive function between patients with ME/CFS and healthy controls. We aimed to determine whether specific cytokine profiles could predict an increased probability of ME/CFS diagnosis as well as poor muscle and cognitive function.

## Materials and Methods

*Study design*. Ninety-two male (n-29) and female (n=63) ME/CFS patients (18-55 years) were recruited through the regional National Health Service CFS services at the Royal Liverpool and Broadgreen University Hospitals, part of the Liverpool University Hospitals Foundation Trust, Merseyside, UK following general practitioner (GP) referral. Patients had an average illness duration of 37 months (range 6-276 months). Ninety-five age- and sex-matched healthy control participants (HCs) were also recruited through local advertisement and via participants family and friends. Participant characteristics are detailed in Table 1. Inclusion and exclusion criteria are described in *SI Appendix* Table S1. All participants provided written informed consent and granted the study team access to their medical notes to obtain medical history. The study was performed in accordance with the Declaration of Helsinki, and the protocol was approved by the University of Liverpool Research Ethics Committee (REC Reference 13/NW/0666; IRAS 115612; February 2014). Two patients with ME/CFS were excluded due to existing diabetes diagnosis and one HC due to insufficient blood sample. The remaining participants provided a blood sample for routine clinical and cytokine analysis (ME/CFS n=90, HC n=94). ME/CFS patients and HCs completed the study questionnaires described below. Sub-groups of participants consented to further optional investigations A) the completion of muscle function testing *in vivo* (ME/CFS n=15, HC n=14) and/or B) provision of a muscle biopsy for immmunohistochemical and mitochondrial function analysis, cytokine and atrogin mRNA analysis and single muscle fibre function measures (ME/CFS n=7-12, HC n=6-9) and/or C) to undergo a battery of cognitive tests (ME/CFS n=12, HC n=12).

**Table 1.**
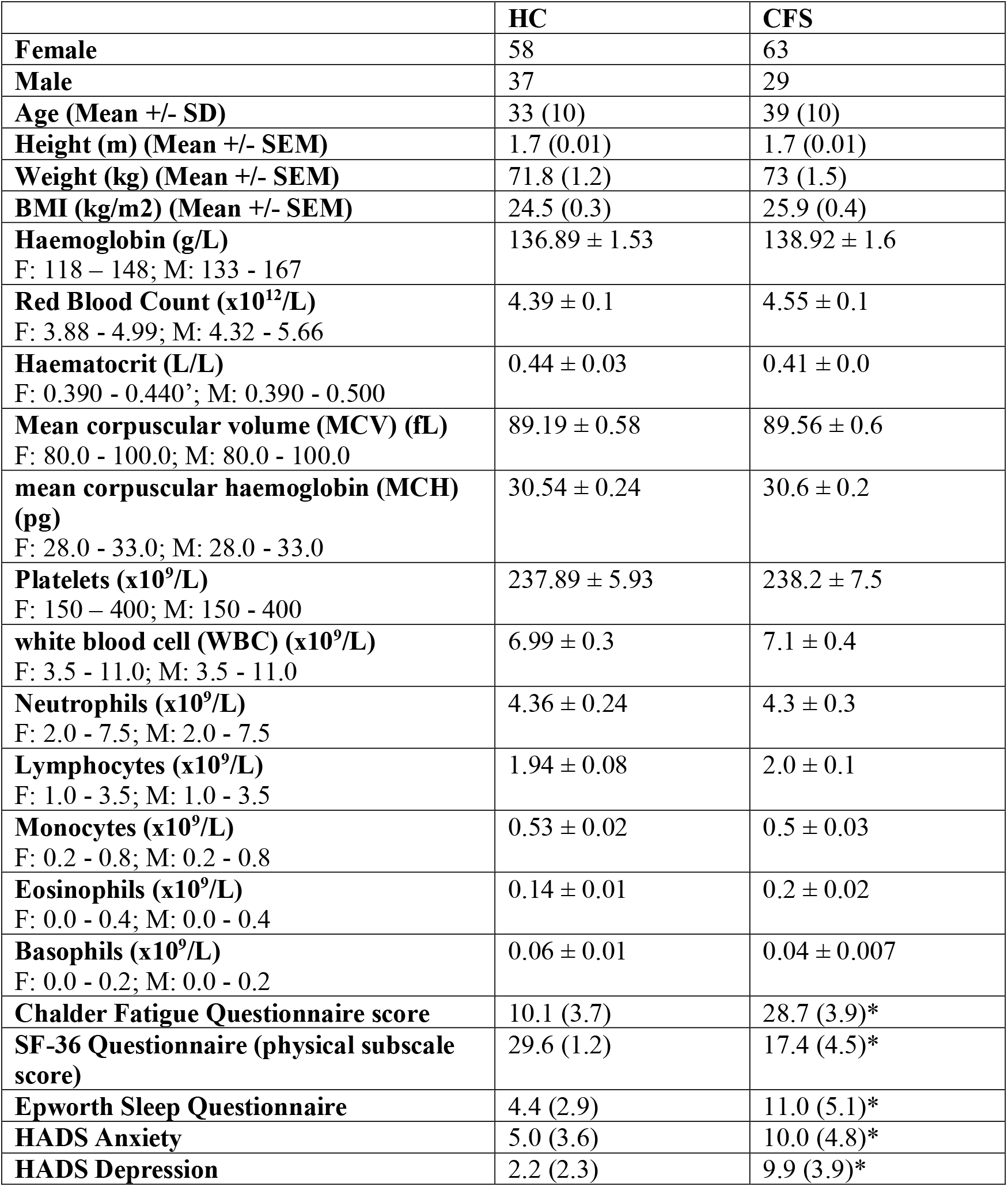
Characteristics of all study participants including routine blood count analysis and questionnaire scores showing normal range values for males (M) and females (F) where appropriate. Patients with ME/CFS: Chalder Fatigue Questionnaire score n=72, SF-36 Questionnaire n=71, Epworth Sleep Questionnaire n=69, HADS Anxiety and Depression n=71. HCs: Chalder Fatigue Questionnaire score n=74, SF-36 Questionnaire n=69, Epworth Sleep Questionnaire n=72, HADS Anxiety and Depression n=74.

Patients with ME/CFS were individually required to complete a series of validated health questionnaires in their Initial Postal Assessment Pack and granted permission for study team to access these questionnaires. These included the Chalder Fatigue Questionnaire, considered a valid and reliable measure of subjective fatigue in patients with ME/CFS (1). Other questionnaires were Hospital Anxiety and Depression Score (HADS), SF-36 (physical subscale) and the Epworth Sleepiness Scale. Healthy controls also completed these questionnaires. Diagnosis of ME/CFS, in accordance with National Institute for Health and Care Excellence (NICE) guidelines (1) was undertaken by clinicians using NICE ME/CFS criteria and Oxford-1991 (45) and CDC-1994/Fukuda (46) criteria. A comprehensive list of symptoms was documented during initial consultation (*SI Appendix* Table S2). Blood was collected for routinely full blood count (FBC) analysis and plasma isolation. Additional blood samples were taken from the sub-group of participants undergoing a muscle biopsy for clotting screen prior to muscle biopsy.

Plasma cytokine concentrations were determined using a 27 cytokine Bio-Plex multi-bead cytokine analysis (PDGF-BB, IL-1β, IL-1Ra, IL-2, IL-4, IL-5, IL-6, IL-7, IL-8, IL-9, IL,10, IL-12, IL-13, IL-15, IL-17, Eotaxin (CCL-11), FGF basic, G-CSF, GM-CSF, IFNγ, IP-10 (CXCL10), MCP-1 (CCL2), MIP1α (CCL3), MIP1 □ (CCL4), RANTES (CCL5), TNF-α, VEGF). Plasma samples were diluted 1 in 4 prior to analysis using the Bio-Plex Pro™ Cytokine Reagent Kit (Bio-Rad, Hercules, USA) and Bioplex® single-plex human cytokine coupled magnetic beads and detection antibodies according to manufacturer’s instructions using the Bioplex® 200 platform (Bio-Rad Hercules, USA).

Muscle function was assessed using a Biodex isometric dynamometer (Biodex Medical Systems Inc. Shirley, NY, USA (47). Participants were seated with a 90° flexion of the hip and non-extendable straps crossing the chest and abdomen and across the quadriceps to maximise isolation of the target muscle group. To assess maximal voluntary contraction (MVC), the angle of knee extension was set at 80° (assuming 0° as full extension). Peak force was the highest level of force achieved during an isometric contraction. To assess involuntary contraction, electrical stimulation of the knee extensors was undertaken using two surface electrodes, (Chattanooga, DJO Global, CA, USA) delivered via a BIOPAC systems MP100 stimulator (BIOPAC systems Inc., Santa Barbara, CA) to generate a force frequency relationship. Fatigue resistance was determined from 2 minutes of repeated electrical stimulation at 30 Hz for 1 sec trains, interspersed by 1 sec relaxation for a total of 60 evoked contractions. The intensity of the applied current was set at an amplitude to elicit 30% of the participants MVC force when stimulated at 100Hz.

A percutaneous needle biopsy was taken from a sub-group of patients with ME/CFS and healthy controls using an aseptic Acecut automatic biopsy system with 22mm needle (TSK Laboratory, Oisterwijk, Netherlands). Multiple passes provided biopsies from the vastus lateralis (VL) of the right leg after skin sterilisation and application of topical antiseptic (chlorhexidine gluconate) prior to the procedure. Muscle biopsies provided samples for histological and immunoblotting analysis, mitochondrial ROS generation and respiration, cytokine mRNA (eotaxin, MIP-1 α, TNF-α, IL-6, IL-8, RANTES, MCP-1, KC and IP-10), Atrogin-1 mRNA content and single muscle fibre functional analysis as previously described (48-50).

Single viable muscle fibres were isolated from the VL muscle biopsies, permeabilized and assessed for contractile properties (48). Fibres were mounted onto an 802D permeabilised fibre apparatus (Aurora Scientific, Canada). Sarcomere length (SL) was measured using 900B Video Sarcomere Length (VSL) software (Aurora Scientific, Canada). Fibres were maximally activated in Ca^2+^ activating solution (pCa 4.5) and peak force was recorded and normalised to fibre CSA (51, 52).

For assessment of mitochondrial ROS generation and respiration, additional muscle biopsy samples were immediately placed into stabilizing buffer. Visible fat and connective tissue were removed and the muscle was manually teased into small fibre bundles. Fibre bundles were permeabilized in preparation for mitochondrial respiration and ROS production analyses of mitochondria in situ (50). Mitochondrial respiration was determined using an oxygen electrode measurement system (Hansatech Instruments Ltd, King’s Lynn, UK) and data standardised to protein content. Respiratory control index (RCI) was calculated by dividing state 3 by state 4 respiration and the efficiency of oxidative phosphorylation was determined by calculating the ratio of ATP amount to consumed O_2_ during state 3 respiration (P:O ratio) (53). Mitochondrial hydrogen peroxide release was measured using Amplex Red-HRP (Molecular Probes, Eugene, OR, USA) as previously described (49, 54, 55). Mitochondrial superoxide generation was determined using a MitoSOX Red, a derivative of dihydroethidium designed for highly selective detection of superoxide in mitochondria (2-OH-Mito-E^+^) with or without the presence of L-NAME to block the activity of nitric oxide synthases (56, 57). Mitochondrial membrane potential (ΔΨm) was assessed using the florescent dye, tetramethylrhodamine, methyl ester (TMRM, 30nM) in response to oligomycin (2.5μM) and the protonophore carbonyl cyanide-p-trifluoromethoxyphenyl hydrazone (FCCP, 4μM), added at the indicated time points. Citrate Synthase activity was determined in muscle homogenates as an index of mitochondrial number using the MitoCheck Citrate Synthase Activity Assay Kit (Cayman Chemical Michigan, USA) and normalised to protein content.

For RNA analysis, muscle biopsies were ground under liquid nitrogen and RNA extracted using Tri Reagent (Sigma Aldrich, Dorset, UK). RNA samples were purified, DNase-treated (RNeasy MinElute cleanup-kit (Qiagen)) and RNA was quantified using a Nanodrop 2000 (Thermo Scientific, Massachusetts, USA). First-strand cDNA was generated from the purified RNA using the iScript cDNA Synthesis kit (Bio-Rad, Hertfordshire, UK). The delta-delta Ct (ΔΔ^Ct^) method was used to quantify changes in gene expression (58). Genes of interest were standardised against housekeeping genes (*SI Appendix* Table S3). Evidence for changes in the production of peroxynitrite by muscle was determined via changes in the level of nitration of tyrosine as previously described (59). Protein carbonyl content of muscle biopsies was assessed using a Rabbit Anti-DNP antibody (Cell Biolabs, San Diego, CA, USA) and evidence of lipid peroxidation was assessed via changes in 4-HNE protein conjugates using rabbit anti 4-HNE (Abcam, Cambridge, UK) primary antibodies (60). Bands were visualized using a Bio-Rad Chemi-Doc System (Bio-Rad Laboratories Ltd, Hemel Hempstead, UK).

A battery of cognitive tests from the Cambridge Automated Neuropsychological Test Battery (CANTAB^®^, Cambridge Cognition UK) were administered individually to ME/CFS and HC on touchscreen laptops. The following cognitive tests were administered and are described in detail in *SI Appendix* Table S4: *Motor Screening Test* (*MOT*)*, Choice Reaction Time* (*RT - at the most difficult level*)*, Paired Associates Learning* (*PAL*)*, Verbal Recognition Memory* (*VRM*)*, Pattern Recognition Memory* (*PRM*). Outcomes subjected to analysis are described in *SI Appendix* Table S4.

### Statistical analysis

All data are presented as mean ± standard error of mean unless stated otherwise. Data were analysed using R v.3.5.0 (61). Data on plasma concentrations of cytokines and muscle function (MVC) were transformed using a logarithmic function to create an approximate normal distribution. Logistic regression was employed to identify cytokines which significantly discriminate ME/CFS patients from HCs controlling for other cytokines in the model. All of the models were adjusted for gender and age. Differences in muscle function between HCs and ME/CFS were investigated using linear regression, adjusted for gender, age, and in subsequent models, relevant cytokines were also included as covariates to examine the relationships between these and muscle function. Model diagnostics were performed after model fitting to assess model suitability. In each model, the best multivariable model was identified as that which had the lowest value of Akaike’s Information Criterion (AIC) (62). AIC is a criterion to identify an optimal combination of predictors in the model; lower AIC indicates better model fit and prediction. Summary scores from the CANTAB tasks were analysed using univariate analysis of variance with age and sex added as covariates or Mann-Whitney tests where there were floor effects. The residuals were examined and where these were not normally distributed, a square root transformation was applied and the analysis rerun. Analysis of reaction times (RT) on the cognitive tests was performed using the Cox proportional hazard model or Cox regression (63). Model fitting was performed using the ‘survival’ package in R (64). The Cox regression is generally employed to fit time-to-event data (65). Individual ‘hazard’, interpreted as the rate at which the participants gave correct answers, is modelled as a function of covariates that modify baseline hazard. The event is defined as a correct answer recorded by the CANTAB software, in which case the reaction time is *uncensored*. When participants make an incorrect response, the reaction time is *censored* since the time to give the correct answer was not observed (65). A higher hazard corresponds to faster speed to give the correct answer, on a given number of trials. In the analysis, we investigated the differences in the hazard between patients with ME/CFS and HCs, adjusted for age, gender, and included cytokines as covariates in subsequent models to assess whether any cytokines significantly predicted RT performance on each cognitive test. Model diagnostics were performed at each modelling stage.

## Results

### Differences between ME/CFS and HC samples in characteristics, symptoms and cytokines

The prevalence of ME/CFS was greater in women in line with previous studies (66). Patients with ME/CFS reported a range of symptoms including post exertional malaise (100%), muscle pain (42%), sleep disturbance (64%) and cognitive dysfunction (29%) (Supplemental Table 2). Data from questionnaire assessments are shown in Table 1. Data demonstrate significantly worse scores by ME/CFS patients on the Chalder Fatigue scale (p-value=0.000) as well as the physical sub-scale of the Short Form (36) Health Survey (SF-36) (p-value=0.000), Epworth Sleep and both HAD scales (largest P<0.001) compared with HCs and adjusted for age and gender, reflecting worse general health status in the patients with ME/CFS compared with HCs. No significant differences were seen in routine blood analyses (Table 1).

### The association of plasma cytokines with diagnosis and symptoms of ME/CFS

Individual plasma cytokine concentrations are shown in Supplemental Figure 1. Only MIP-1β differed significantly (p=0.044) between HC and ME/CFS, after controlling for age and sex. However, such bivariate correlations, do not take into account relationships between cytokines. The results of an optimised multivariate logistic regression analysis, where CFS/HC status is regressed on the cytokines *simultaneously* are presented in Table 2 and the directional changes in cytokine concentrations associated with increased disease are summarised in *SI Appendix* Table S5. The model showed that PDGF, IL-6, IL-10, eotaxin, MIP-1α, MIP-1β, RANTES, and VEGF were significantly associated with diagnosis (HC or ME/CFS) after adjusting for age and gender, indicating that diagnosis of ME/CFS is significantly associated with increased plasma content of IL-10, MIP-1β, and RANTES and lower PDGF, IL-6, eotaxin, MIP-1α and VEGF. In this analysis, CFS/HC status was also marginally associated with increased IP-10 (p=0.053). When individual associations between Chalder Fatigue scores and plasma cytokines, adjusted for age and gender, were examined only MIP-1α and IP-10 show significant associations. However, when taking into account the influence of correlations between cytokines by entering them into a joint model (Table 3 and *SI Appendix* Table S5), higher plasma values of IL-1β, IL-8, IL-10, IP-10, and RANTES were significantly associated with higher Chalder Fatigue scores i.e. greater perceived fatigue, after adjusting for age and gender and taking into account the influence of other cytokines in the mode and lower plasma values of PDGF, eotaxin, G-CSF, and MIP-1α were associated with greater perceived fatigue. Similarly, lower values of PDGF and MIP-1a and higher values of IL-10 and RANTES were associated with lower SF-36 scores (worse physical health) (Table 4 and *SI Appendix* Table S5).

**Table 2.** Association between CFS/HC status and plasma cytokine concentrations, adjusted for Age+Sex, in logistic regression after variable selection (optimal model). Estimates, standard errors, test statistic (z-value), and statistical significance (p-value) of predictors in the optimal multivariable logistic regression in identifying significant factors to discriminate ME/CFS from healthy controls. The asterisks (*) indicate p-values less than 0.05.

| **Predictor** | **Estimate** | **Standard error** | **Z-value** | **p-value** |
| --- | --- | --- | --- | --- |
| **(Intercept)** | -13.2653 | 4.3177 | -3.07 | 0.0021* |
| **Age** | 0.0697 | 0.0197 | 3.54 | 0.0004* |
| **Gender** | -0.6627 | 0.4091 | -1.62 | 0.1053 |
| **RANTES** | 1.3876 | 0.386 | 3.59 | 0.0003* |
| **MIP-1α** | -0.7357 | 0.2563 | -2.87 | 0.0041* |
| **Eotaxin** | -1.6608 | 0.6149 | -2.70 | 0.0069* |
| **IL-10** | 0.6061 | 0.2448 | 2.48 | 0.0133* |
| **PDGF-bb** | -0.4069 | 0.1753 | -2.32 | 0.0202* |
| **MIP-1β** | 1.172 | 0.54 | 2.17 | 0.0300* |
| **IL-6** | -0.7677 | 0.377 | -2.04 | 0.0417* |
| **VEGF** | -0.1762 | 0.0895 | -1.97 | 0.0490* |
| **IP-10** | 0.8549 | 0.4409 | 1.94 | 0.0525 |
| **IL-13** | 0.4721 | 0.2712 | 1.74 | 0.0817 |

**Table 3.** Estimates, standard errors, test statistic (t-value), and statistical significance (p-value) of predictors in the optimal multivariable linear regression in which Chalder Fatigue Score is the outcome predicted. The asterisks (*) indicate p-values less than 0.05.

| **Covariates** | **Estimate** | **Std. Error** | **t value** | **p-value** |
| --- | --- | --- | --- | --- |
| **(Intercept)** | 5.0705 | 18.8293 | 0.27 | 0.7882 |
| **Age** | 0.4271 | 0.0776 | 5.51 | 0.0000* |
| **Gender** | -3.8356 | 1.5483 | -2.48 | 0.0146* |
| **MIP.1a** | -3.6842 | 0.8816 | -4.18 | 0.0001* |
| **IL.10** | 2.8758 | 0.9063 | 3.17 | 0.0019* |
| **IL.8** | 9.1933 | 3.011 | 3.05 | 0.0028* |
| **IL.1b** | 3.7972 | 1.4853 | 2.56 | 0.0118* |
| **IP.10** | 4.2833 | 1.7016 | 2.52 | 0.0131* |
| **PDGF** | -1.7933 | 0.7397 | -2.42 | 0.0168* |
| **RANTES** | 3.5628 | 1.4751 | 2.42 | 0.0172* |
| **Eotaxin** | -6.2552 | 2.8633 | -2.18 | 0.0308* |
| **G.CSF** | -5.0902 | 2.3911 | -2.13 | 0.0352* |
| **IL.6** | -3.0356 | 1.5841 | -1.92 | 0.0576 |
| **IL.13** | 2.0711 | 1.0892 | 1.9 | 0.0596 |
| **IL.1ra** | -1.8337 | 0.9847 | -1.86 | 0.0649 |
| **IL.4** | -2.9141 | 1.5794 | -1.85 | 0.0674 |
| **IL.7** | -3.1519 | 2.0857 | -1.51 | 0.1333 |

**Table 4.**
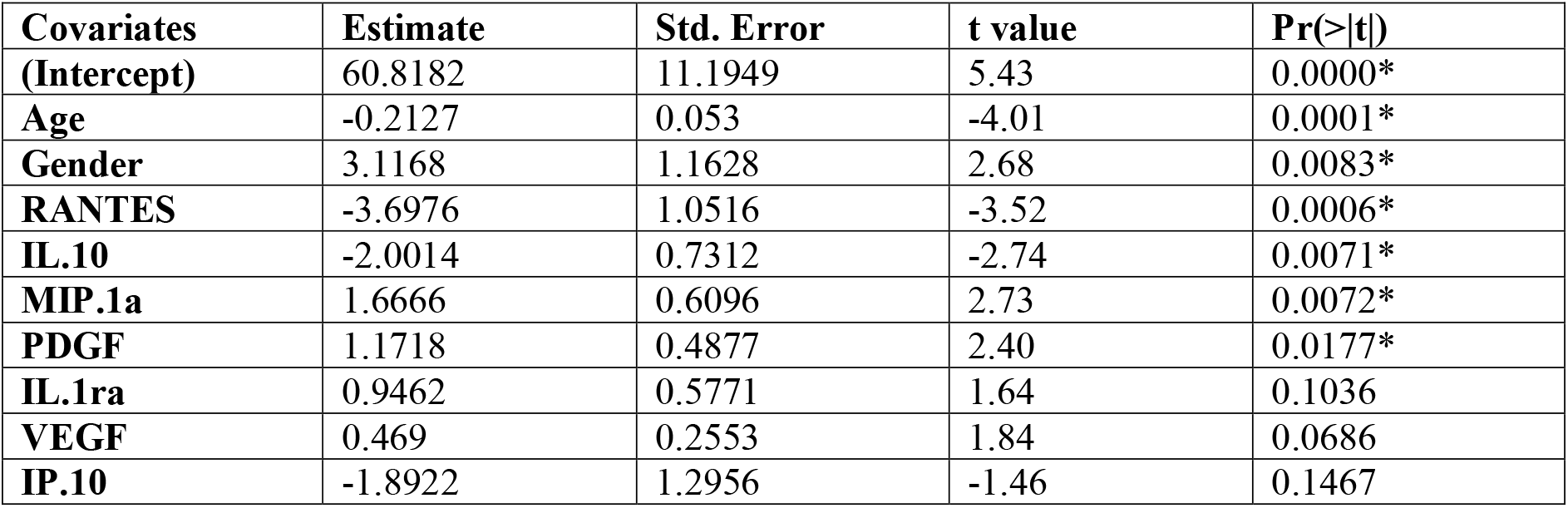
Estimates, standard errors, test statistic (t-value), and statistical significance (p-value) of predictors in the optimal multivariable linear regression in which SF-36 Score is the outcome predicted. The asterisks (*) indicate p-values less than 0.05.

| **Covariates** | **Estimate** | **Std. Error** | **t value** | **Pr(>\|t\|)** |
| --- | --- | --- | --- | --- |
| **(Intercept)** | 60.8182 | 11.1949 | 5.43 | 0.0000* |
| **Age** | -0.2127 | 0.053 | -4.01 | 0.0001* |
| **Gender** | 3.1168 | 1.1628 | 2.68 | 0.0083* |
| **RANTES** | -3.6976 | 1.0516 | -3.52 | 0.0006* |
| **IL.10** | -2.0014 | 0.7312 | -2.74 | 0.0071* |
| **MIP.1a** | 1.6666 | 0.6096 | 2.73 | 0.0072* |
| **PDGF** | 1.1718 | 0.4877 | 2.40 | 0.0177* |
| **IL.1ra** | 0.9462 | 0.5771 | 1.64 | 0.1036 |
| **VEGF** | 0.469 | 0.2553 | 1.84 | 0.0686 |
| **IP.10** | -1.8922 | 1.2956 | -1.46 | 0.1467 |

**Table 5.** Estimates, standard errors, test statistic (t-value), and statistical significance (p-value) of predictors in the optimal multivariable linear regression in identifying cytokines which significantly predict maximal voluntary contraction (MVC) in log scale. The asterisks (*) indicate p-values less than 0.05.

|  | **Estimates** | **Standard Error** | **t-value** | **p-value** |
| --- | --- | --- | --- | --- |
| **(Intercept)** | 7.8958 | 0.9741 | 8.11 | 0.0000* |
| **Gender** | 0.1269 | 0.1004 | 1.26 | 0.2245 |
| **IL.8** | -1.225 | 0.1879 | -6.52 | 0.0000* |
| **IP.10** | -0.8921 | 0.1633 | -5.46 | 0.0001* |
| **Eotaxin** | 0.8003 | 0.1467 | 5.46 | 0.0001* |
| **TNF.a** | -0.3738 | 0.0761 | -4.91 | 0.0002* |
| **IL-1β.** | 0.3523 | 0.0796 | 4.42 | 0.0004* |
| **PDGF** | 0.1635 | 0.0378 | 4.32 | 0.0005* |
| **G.CSF** | 0.5586 | 0.142 | 3.93 | 0.0012* |
| **IL.13** | 0.0933 | 0.0586 | 1.59 | 0.1312 |
| **IL.1ra** | 0.1014 | 0.0821 | 1.24 | 0.2344 |
| **MIP.1b** | -0.1996 | 0.168 | -1.19 | 0.2520 |
| **IL.7** | 0.1246 | 0.1158 | 1.08 | 0.2976 |

### Muscle function in ME/CFS and association with cytokines

Maximal voluntary contraction (MVC) force was significantly reduced in patients with ME/CFS compared with HCs (Figure 1A; p=0.0272, after adjusting for age and gender). In contrast, there was no significant difference in the generation of maximal involuntary contraction by electrical stimulation between the two groups overall and at any frequency (Figure 1B). Figure 1C indicates that the rate of loss of torque (rate of fatigue during electrical stimulation to contract) appeared greater in the muscle of patients with ME/CFS compared with HCs and this difference increased with duration of stimulation. However, there was no significant difference between HC and ME/CFS and no group by time interaction. Histological analysis of muscle fibres demonstrated no overt pathology, but fibre size analysis showed a significantly greater percentage of muscle fibres below 50um in diameter in patients with ME/CFS (Figure 1D). Analysis of maximum contractile force generation (activated by calcium) by isolated permeablized single fibres showed no difference between patients with ME/CFS and HCs (Figure 1E) including when normalised to fibre size (Figure 1F). A linear regression model to identify cytokines that significantly predict lower MVC taking into account the influence of other cytokines indicated that higher values of IL-8, IP-10, and TNF-α and lower values of PDGF, IL-1β, eotaxin, and G-CSF are associated with lower MVC. The association of such directional changes in cytokine concentration on poor MVC is reflected in *SI Appendix* Figure S5.

**Figure 1.**
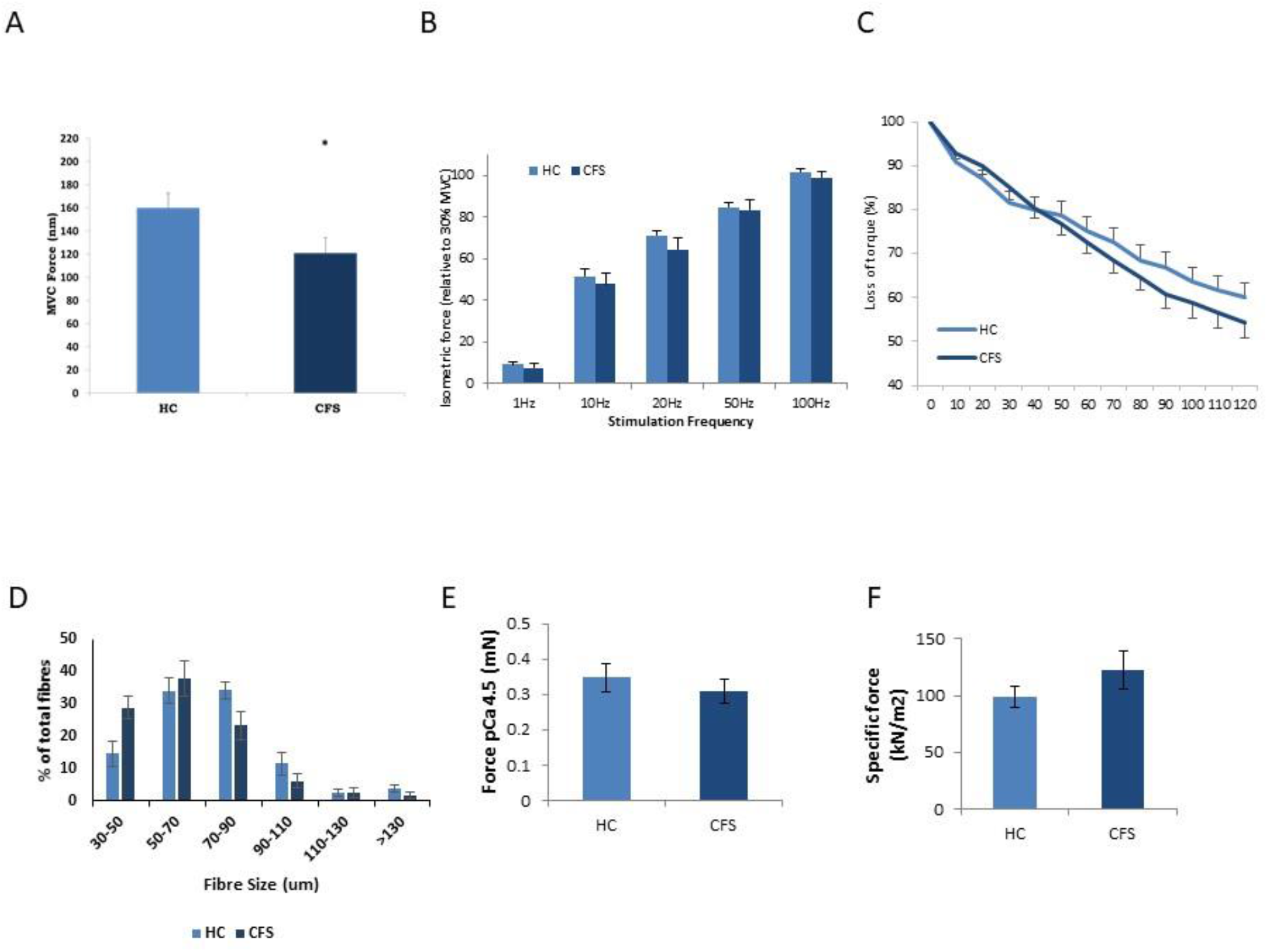
**(A)**. Maximal voluntary isometric force of the right knee extensors in HCs and patients with ME/CFS The asterisk (*) indicate p-values less than 0.05; (**B**) Maximal involuntary contraction, electrically stimulated force frequency relationship of the right knee extensors in HCs and patients with ME/CFS; (**C**) Fatigue of the right knee extensors in HCs and patients with ME/CFS when subjected to 120 seconds of repeated 1 second, 30 Hz trains interspersed with 1 second rest; (**D**) Histological analysis of muscle fibre sizes (**E**) Maximum Ca^2+^ activated tetanic force of isolated single muscle fibres from patients with ME/CFS and HCs. (**F)** Maximum Ca^2+^ activated force in isolated single muscle fibres normalised to fibre cross-sectional area in HCs and patients with ME/CFS.

## Analysis of muscle biopsy samples

### Analysis of muscle samples for mitochondrial ROS production, respiration and mitochondrial membrane potential were performed in a subsample of ME/CFS patients (n=7) and HCs (n=6)

Figure 2A shows changes in 2-OH-Mito-E^+^ fluorescence over time in muscle fibres from patients with ME/CFS and HCs. Data demonstrated no significant difference in mitochondrial superoxide generated by muscle fibres from patients with ME/CFS and HCs either prior to (Figure 2A) or following (Figure 2B) the addition of the nitric oxide synthetase inhibitor L-NAME. No difference in mitochondrial H_2_O_2_ generation was seen either with endogenous or added substrates in muscle bundles from patients with ME/CFS compared with HCs (Figure 2C). Data for citrate synthase activity, mitochondrial respiration, P:O ratio demonstrated no significant changes in mitochondrial number (represented by citrate synthase activity), resting mitochondrial respiration or efficiency of oxidative phosphorylation. (*SI Appendix*, Figure 2). Similarly, no significant differences in mitochondrial membrane potential were seen in response to oligomycin and the protonophore FCCP oxidative phosphorylation inhibitors (Figure 2D). No evidence for changes in oxidative damage were observed between patients with ME/CFS and HCs, shown by the analyses of protein oxidation, lipid peroxidation and the levels in protein nitration (3-NT) (data not shown). Representative images for MitoSOX Red and TMRM analyses are shown in *SI Appendix* Figure S3.

**Figure 2.**
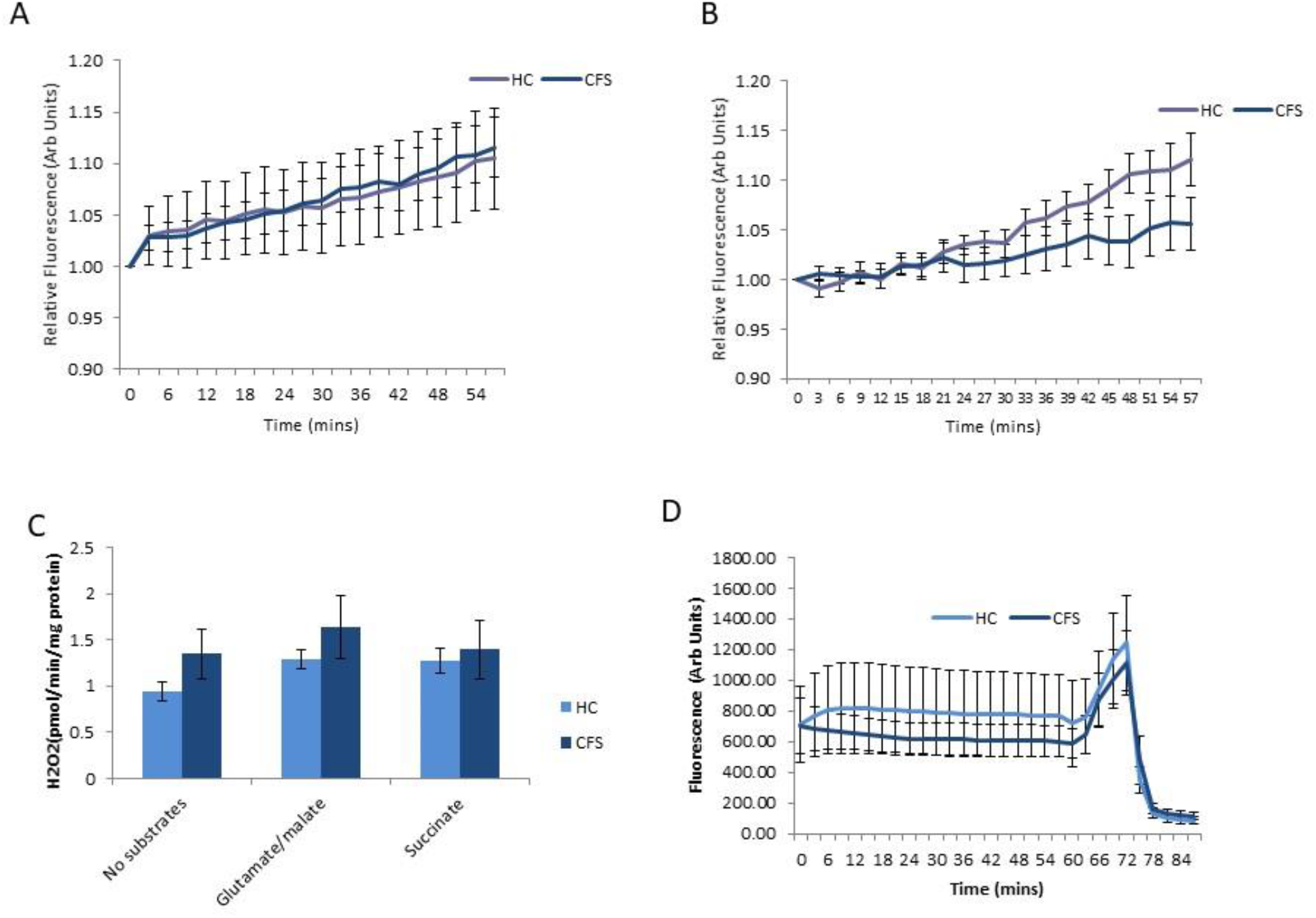
Changes in mitochondrial 2-hydroxyethidium (2-OH-Mito-E^+^) fluorescence assessed in permeabilised fibre bundles prepared from *vastus lateralis* muscle of patients with ME/CFS and HCs, without **(A)** and with **(B)** L-Name; **(C)** Generation of mitochondrial H_2_O_2_ assessed in permeabilised fibre bundles prepared from *vastus lateralis* muscle of patients with ME/CFS and HCs in the presence of mitochondrial substrates and inhibitors, Glutamate/Malate and Succinate and normalised to protein content of the muscle; **(D)** Mitochondrial membrane potential (ΔΨm) in intact mitochondria of isolated *vastus lateralis* fibres bundles from patients with ME/CFS and HCs assessed by changes in TMRM fluorescence in response to oligomycin (Olm) and the protonophore (FCCP), added at the indicated time points. Data presented as mean ± SEM.

### Cytokine and atrogin mRNA content of muscle biopsies

Muscle biopsies were analysed for mRNA content of a number of cytokines (eotaxin 1, MIP-1α, TNF-α, IL-6, IL-8, RANTES, MCP-1, KC and IP-10), however, significant increases were only detected in mRNA content of MCP-1 and IP-10 suggesting the specific increase in production of these cytokines by skeletal muscles of patients with ME/CFS (Figure 3). Levels of Atrogin 1 mRNA were also significantly elevated in muscle biopsies of patients with ME/CFS compared with HCs (Figure 3) suggesting an increase in protein degradation processes in skeletal muscles of patients with ME/CFS compared with HCs.

**Figure 3.**
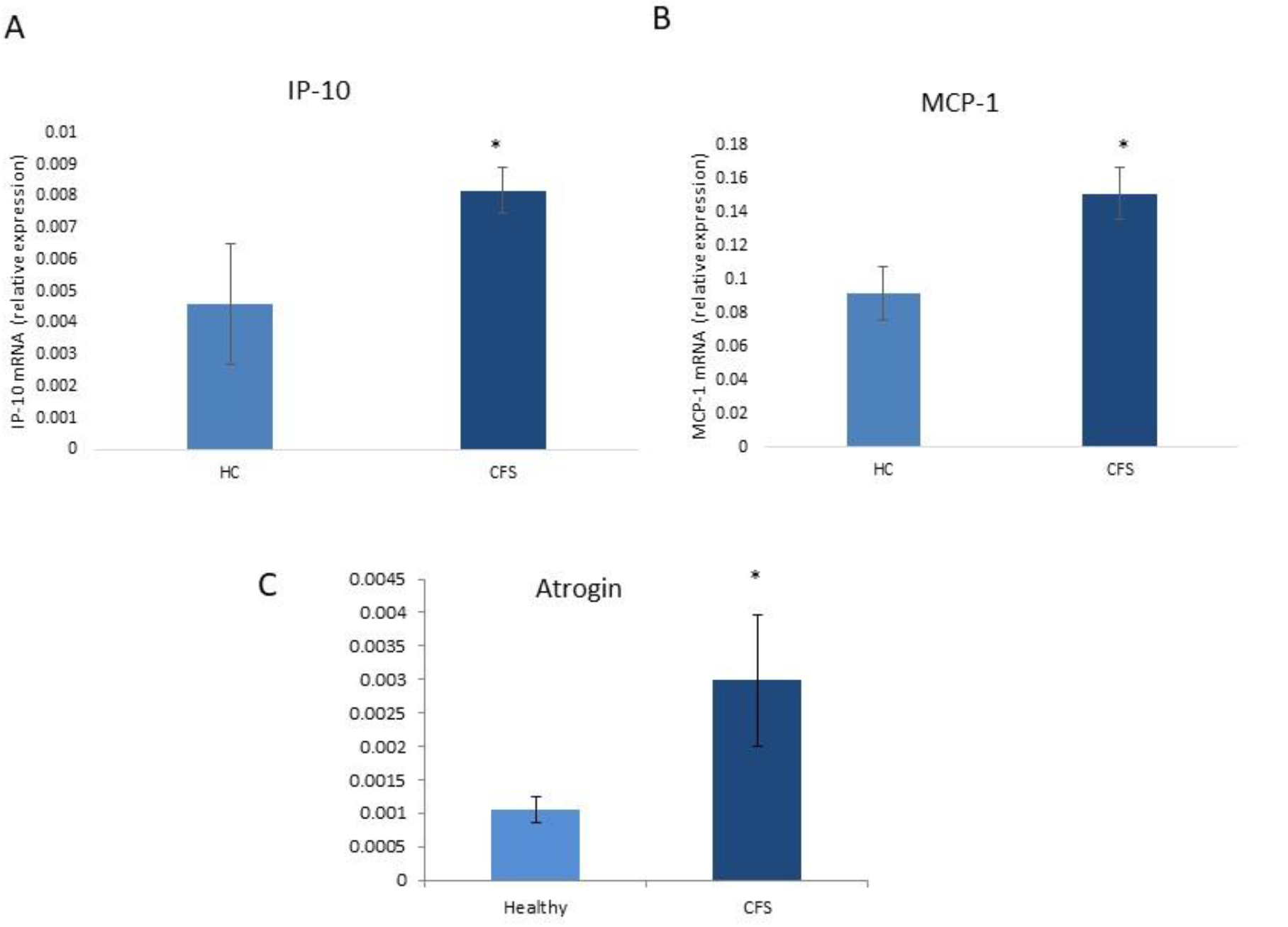
Relative mRNA expression of IP-10, MCP-1 and Atrogin in muscle homogenates from patients with ME/CFS and HCs. Data are presented as fold change and presented as mean ± SEM. The asterisks (*) indicate p-values less than 0.05.

## Cognitive function outcomes

Cox regression analysis was performed to identify significant factors that affect the ‘hazard’ (rate of making a correct response at any time) for all cognitive outcomes (Figure 4). Without adjusting for cytokines, the results indicate that patients with ME/CFS were more likely to have slower RTs than HCs on the immediate PRM task (Chi-sq(1)=6.09, p value=0.0136) (shown in Figure 4) and immediate VRM task (Chi-sq(1)=61.87, p-value=0.000), but not on MOT, VRM Recall, nor delayed PRM. There were no significant differences between patients with ME/CFS and HCs on any accuracy or error outcomes from any of the cognitive tests (*Motor Screening Test* (*MOT*)*, Choice Reaction time* (*RT*)*, Paired Associates Learning* (*PAL*)*, Pattern Recognition Memory* (*PRM*) with the exception of *Verbal Recognition* Memory (VRM) (*SI Appendix* Table S6), where patients with ME/CFS recalled significantly fewer words than HCs (F(1,23)=6.88, p=0.016).

**Figure 4.**
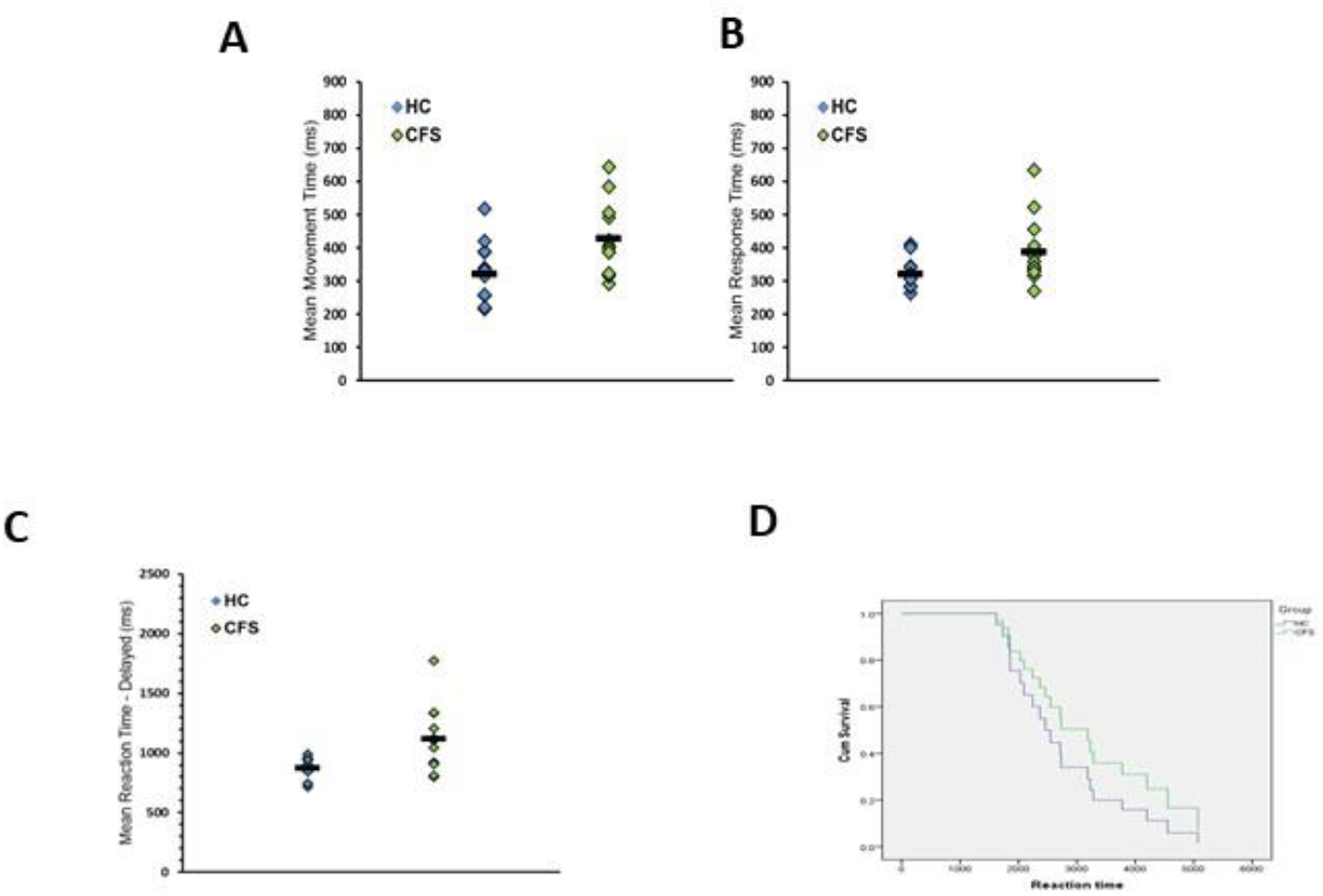
Significantly slower **(A)** movement and **(B)** total RT in patients with ME/CFS compared with HCs on the 5 Choice RT task. Univariate analyses, P<0.05 Bar =mean RT. **(C)** Delayed Pattern Recognition Memory (PRM-del) **(C)** RT and **(D)** Survival Plot indicating that patients with ME/CFS had lower probability of responding quickly than HCs at any given time - B= −0.45 (0.14), p=.001, Exp(B)=0.64 (95% CI = 0.49-0.83).

### Motor Screening Test (MOT)

For Reaction Time (RT) on the motor screening task (MOT), there was a significant difference between patients with ME/CFS and HCs after adjusting for age, gender with plasma cytokines included as covariates, (Chi-sq=8.68, p-value=0.0032). Results indicate that HCs were, on average, faster to make a correct response than patients with ME/CFS and that a slower RT was associated with higher levels of IL-5, VEGF, IL1Rα, IFNγ, eotaxin, IL-12 and G-CSF. Higher levels of TNFα, IL-4, and IL-10, were associated with faster responding and therefore increased likelihood of being a HC. Because it is possible that differences in RT between ME/CFS and HCs were due to changes in motor function, i.e. volitional muscle function, we related our measure of muscle function and fatigue to RT from the cognitive tests by including MVC as a covariate in the analysis of RTs for each cognitive outcome. Time to respond correctly on MOT was significantly associated with MVC after adjusting for age and gender (Chi-sq=8.66, p-value=0.0033) and remained significant when adjusted for plasma cytokines (Chi-sq=6.07, p-value=0.0137). Slower RT was associated with higher levels of IL1ra, IL-5, IL-12 and IP10 and lower levels of IL-9, MIP-1β and TNFα. Chalder Fatigue score was also significantly associated with RT after adjusting for age and gender (Chi-sq=8.91, p-value=0.0028) and plasma cytokines (Chi-sq=22.57, p-value=0.000). Specifically, lower levels of PDGF, IL-5, IL-10, MIP-1β and RANTES were associated with slower responses as were higher levels of IL-4, IL-9 and VEGF. Score on SF-36 was not associated with RT when adjusted for age and gender. Inclusion of plasma cytokines resulted in a significant association between SF36 and MOT performance (Chi-sq=7.07, p-value=0.0078). Here, slower RT was associated with lower levels of IL1ra, IL-5, IL-10, IP-10 and MIP-1α and higher levels of IL-4, IL-9, and MIP-1β. Data are summarised in *SI Appendix* Table S7A-D.

### Choice Reaction Time (CRT)

For choice reaction time, we analysed the harder trials where a choice 5 stimuli was required and controlled for age and sex in each model. Patients with ME/CFS were significantly slower to make a correct response than HCs (Chi-sq=33.17, p<0.0001). Slower RT was associated with higher IL-13, RANTES, FGF, IL-17, IL-1β, IL-12, I-L4, and TNFα and increased likelihood of ME/CFS. Higher levels of MIP-1α, MIP-1β, PDGF, IL-5, IL-9, IL-10, IL-1Ra, IL-8 and IL-7 were associated with faster RT and being a HC. MVC was associated with RT after adjusting for age and gender (Chi-sq = 12.50, p=0.0004 respectively) but MVC was no longer significant when adjustment for cytokines was made. Slower RT was associated with higher IFNγ, IL-5, IL-1β and faster RT with higher levels of TNFα and IL-4. Chalder fatigue score was associated with RT, after adjusting for age and gender, (Chi-sq= 34.92, p<0.0001) and remained significantly associated when adjusted for plasma cytokines (Chi-sq = 25.31, p=0.002). Higher MIP-1β, IL-8, IL-9, IFNγ and IL-5 were associated with slower RT whereas faster RT was associated with higher levels of IL-13, RANTES, FGF, MIP-1P, PDGF and IL-1β. SF36 was associated with RT after adjusting for age and gender (Chi-sq = 18.75, p>0.0001) and remained significantly associated with RT when cytokines were included (Chi-sq= 27.01, p=0.0001). Higher IL-13, MIP-1α, IL-1Ra, and IP-10 were associated with slower and MIP-1β and IL-8 with faster RT. Data are summarised in *SI Appendix* Table S7A-D.

### Pattern recognition memory (PRM)

Pattern recognition memory (PRM) RT assessed immediately after presentation showed a difference between patients with ME/CFS and HCs in time to make a correct response after adjusting for age and gender (Chi-sq=6.09, p-value=0.0136). When plasma cytokines were included as covariates, this difference between ME/CFS and HC remained significant (Chi-sq=5.56, p-value=0.0184). Thus, for PRM HCs responded faster than patients with ME/CFS. Slow RT was significantly associated with higher levels of VEGF, IFNγ, IL-12, IL-9, IL-6, G-CSF, MIP-1β, and eotaxin, whereas IL-10, IL-17, FGF, IP-10, IL-1Ra, IL-13, IL-8, TNFα, and MIP1α were associated with faster responses. MVC was significantly associated with response speed on PRM after adjusting for age and gender (Chi-sq=13.0, p-value=0.0003). However, adding plasma cytokines rendered the association non-significant. Nevertheless, lower levels of IL-4, IL-12, and FGF were found to be significantly associated with slower RT and higher levels of IL-5, IL-8, and IL-17 were associated with slower RT on the immediate pattern recognition memory task. Chalder Fatigue score was also significantly associated with RT on the PRM task (Chi-sq=9.47, p-value=0.0021), and remained so when adjusted for plasma cytokines (Chi-sq=5.56, p-value=0.0184). Lower values of IL-6, IL-12, Eotaxin, GCSF, IFNγ, MIP-1β, and VEGF were associated with slower PRM immediate RT, while higher IL-8, IL −13, IL-17, IP-10, and TNFα were associated with slower RT. SF-36 score was significantly associated with PRM RT, when adjusted for age and gender (Chi-sq=5.27, p-value=0.0217), but the association between MVC and PRM RT was not significant when plasma cytokines were added to the model. Nevertheless, lower levels of IL-6, FGF, and MIP-1β and higher levels of IL-13 and IL-17 were found to be associated with slower RT on the PRM immediate task. Data are summarised in *SI Appendix* Table S7A-D.

### Delayed Pattern recognition memory (PRM)

Delayed Pattern recognition memory (PRM-del) RT was also assessed after a delay of 30 minutes following presentation. The difference between patients with ME/CFS and HCs in time to make a correct response after adjusting for age and gender failed to reach significance (Chi-sq= 2.73, p-value=0.0988). MVC was not significantly associated with PRM-del RT (Chi-sq=2.58, p-value=0.1085). Adjusting for plasma cytokines, the association became significant (Chi-sq=4.89, p-value=0.0270). Lower values of IP-10 and MIP1α and higher values of IL-13 and IL-6 were associated with slower PRM-del RT. Chalder Fatigue score was also significantly associated with PRM-del RT, after adjusting for age and gender (Chi-sq=5.43, p-value=0.0198) but adjusting for plasma cytokines rendered the association non-significant. SF36 score failed to reach significance, after adjusting for age and gender, (Chi-sq= 3.50, p-value=0.0613) and remained non-significant after adjusting for plasma cytokines. Data are summarised in *SI Appendix* Table S7A-D.

### Immediate verbal recognition memory (VRM)

VRM-RT indicated a highly significant difference between patients with ME/CFS and HCs after adjusting for age and gender (Chi-sq=61.87, p-value=0.0000), with HCs considerably faster than patients with ME/CFS. Inclusion of cytokines in the model removed this significant difference, which suggests that the cytokines explain the variance associated with variability in RT. Slower responses times were significantly associated with higher IL-17, IP-10, IL-13 and lower VEGF, MIP-1α, and IL-7. MVC was found to be significantly associated with VRM RT after adjusting for age and gender (Chi-sq=20.91, p-value=0.0000). After adjusting for plasma cytokines, this association became non-significant. Slower response times were significantly associated with higher PDGF, IL-1α, IL-6, IL-8, IL-13, TNFα, VEGF and faster RT with IL-1β, IL-12, IL-17, IFNγ, MIP-1β and RANTES. Response speed on verbal recognition memory (VRM), was significantly associated with MVC, Chalder Fatigue score, and SF36 after adjusting for age and gender (MVC: Chi-sq=20.91, p-value=0.000; Chalder Fatigue score: Chi-sq=48.92, p-value=0.000; SF36: Chi-sq=43.45, p-value=0.000). However, all of these associations became non-significant when adjusted for plasma cytokines. Data are summarised in *SI Appendix* Table S7A-D.

For VRM delayed recall, there was no difference in the RT between patients with ME/CFS and HC groups and adjustment for age, gender and cytokines did not improve discrimination. Nevertheless, lower levels of PDGF, IL-1β, IL-1Ra, IL-12, Eotaxin, FGF, GCSF and RANTES were associated with slower VRM delayed recall. Higher values of IL-7, IL-8, IL-9, IL-10, IL-17, and TNFα were associated with slower VRM delayed recall. MVC was not associated with VRM delayed recall when adjusted for age and gender. The association was significant when it was further adjusted with plasma cytokines (Chi-sq=4.51, p-value=0.0338). Higher values of IL-7 and IL-12 were associated with slower RT. Chalder fatigue score was not associated with VRM delayed recall after adjusting for age and gender but inclusion of plasma cytokines in the model resulted in significant associations with response speed. (Chi-sq=7.64, p-value=0.0057). In this model, lower levels of PDGF, IL-1Ra, IL-5, IL-12, Eotaxin, and GCSF are associated with slower VRM delayed recall, while higher levels of IL-7, IL-8, IL-9, IL-10, IL-13, IL-17, FGF, MIP-1β, TNFα, and VEGF were associated with slower RT. SF36 score was not associated with response speed irrespective of covariates in the model. Higher levels of PDGF, IL-10, and FGF were associated with faster VRM delayed recall while higher levels of IL-1ra and IL-12 were associated with slower RT. Data are summarised in *SI Appendix* Table S7A-D.

## Discussion

The purpose of the current study was to comprehensively assess muscle function and plasma immune profiles in patients with ME/CFS and is the first study to use newly developed techniques to permit examination of skeletal muscle mitochondrial function with a novel *in situ* approach. The study further investigated the potential contribution of muscle-derived cytokines to the plasma immune profile for both healthy control and age matched ME/CFS patients. Our analysis of individual plasma cytokines indicated that, with the exception of MIP-1β, no cytokine was singularly associated with ME/CFS or HC status, after adjustment for age and gender. Cytokines do not act alone, can have pleiotrophic actions and interact in pathways. In this analysis, we have taken into account the dependencies between plasma cytokines by considering a joint model for prediction of ME/CFS. This detailed and novel analysis has clearly identified that newly diagnosed patients with ME/CFS demonstrated distinguishing differences in certain plasma cytokines, when adjusted for age, gender and that cytokine interactions predicted both diagnosis and a number of measures common to patients with ME/CFS including MVC and perceived fatigue as well as cognitive indices such as pattern and verbal reaction times.

### Muscle function and fatigue in patients with ME/CFS

Extensive examination of muscle function was undertaken in this study. The patients with ME/CFS demonstrated a characteristic reduction in MVC (Figure 1A). The reduced MVC and increased perception of fatigue in patients with ME/CFS compared with HCs was reflected in the health questionnaires. In contrast, the maximum contractile forces generated by electrical stimulation of lower limb muscles of patients with ME/CFS were similar to those of HCs (Figure 1B) as previously reported (67). Decreased function causing failure of voluntary muscle contraction can occur at all levels of the neuromuscular system, including the motor cortex, signaling to motoneurons, neurotransmitter and motoneuron signals to the muscle, excitation-contraction coupling in the muscle, and actin-myosin filament interactions (67, 68). The lack of difference in electrically stimulated contractions was reflected in similar contractile force generation by isolated skinned muscle fibres, activated by increasing calcium concentrations, suggesting no defect in the actin-myosin cross-bridge function in muscles of patients with ME/CFS. Electrical stimulation of muscles to contract using surface (skin) electrodes removed the need for central and peripheral neural input hence the data suggest that there was no major defect evident in the post-synaptic activation of the muscles, at least for a single maximal contraction. No significant difference was seen in the rate of fatigue of muscles when electrically stimulated to contract in patients with ME/CFS (Figure 1C), also suggesting no intrinsic defect in contractile proteins. Levels of Atrogin 1 mRNA were significantly elevated in muscle biopsies of patients with ME/CFS compared with HCs suggesting an increase in protein degradation processes in muscles of patients with ME/CFS compared with HCs. Atrogin-1 binds to polyubiquitinated proteins to direct them for subsequent degradation by the 26S proteasome and as such is an important regulator of ubiquitin-mediated protein degradation in skeletal muscle (69). Increased levels of Atrogin-1 mRNA are associated with reduced muscle mass (69) and in this study Atrogin-1 was associated with a significant reduction in muscle fibre size, although a detailed examination of muscle protein degradation was not undertaken.

### Mitochondrial function and ME/CFS

Defective mitochondria have been implicated in conditions where fatigue is prevalent such as ME/CFS, but whether mitochondria are abnormal in patients with ME/CFS remains the subject of considerable debate. Contrasting findings are likely to be due to a number of factors, particularly the limited methods that have traditionally involved isolation of mitochondria for subsequent study, although the validity of this approach has recently been questioned (70). To overcome the limitations of approaches involving mitochondrial isolation, a method of isolating bundles of muscle fibres and permeabilization with saponin has been employed in this study. Electron microscopic examination of isolated fibres demonstrated that the mitochondria remain intact in this model (71) and so this approach allowed examination of function of mitochondria that remain in their original location within fibres. The mechanisms by which mitochondrial abnormalities can result in an altered immune profile are poorly understood, although several studies have demonstrated that dysfunctional or failing mitochondria can release mitochondrial-derived damage –associated molecular patterns (72) and that these are associated with chronic inflammation in conditions such as ageing and several degenerative diseases. Inflammation can occur in the absence of infection and it has been proposed that mitochondria contribute to such ‘sterile inflammation’(73). No difference in mitochondrial superoxide or hydrogen peroxide generation by mitochondria of patients with ME/CFS was seen compared with HCs, either in State 1 (with endogenous substrates) or with the addition of glutamate/maleate or succinate. This lack of difference could not be explained by changes in absolute numbers of mitochondria since data showed similar levels of citrate synthase activity in fibres from patients with ME/CFS. The lack of difference in mitochondrial membrane potential between the HC and ME/CFS samples also suggests no major structural abnormalities in muscle mitochondria. A functional increase in mitochondrial oxidative stress and hydrogen peroxide production would be predicted to result in increased oxidative damage to skeletal muscle although little evidence of this was seen in muscles of patients with ME/CFS compared with HCs in a similar manner to other studies (29, 30). Dysfunctional mitochondria have been reported in non-muscle cells from patients with ME/CFS (eg (28)) and so although the current study suggests that there are no major mitochondrial abnormalities in skeletal muscle mitochondria, it does not rule out the potential role of dysfunctional non-muscle mitochondria.

### Cognitive function and ME/CFS

Data demonstrated significant increases in RTs in all of the cognitive function tests employed although no significant difference in accuracy was seen. The slower reaction times were significantly correlated with MVC, suggesting a role for motor function in the slower RTs in patients with ME/CFS. Data also demonstrated that the differences in RTs between ME/CFS and HC were in cognitive measures that are immediate. This difference was not evident in the delayed measures. Twenty nine percent of the patients with ME/CFS in this study reported aspects of cognitive disfunction. Such changes have been well characterised in a number of other studies where patients have reported deficits including poor selective and divided attention, slow information processing and vulnerability to distraction (74). In some studies, cognitive performance was inversely correlated with exertion and fatigue (74) and this was evident in the current study, where time to respond correctly in several cognitive tests were significantly associated with MVC and Chalder Fatigue measures.

### Identification of cytokine patterns predictive of ME/CFS

Many patients with ME/CFS report having experienced a viral or bacterial infection directly prior to the onset of their illness (8-12). This has led researchers to investigate the hypothesis that inflammation may be a mechanism by which ME/CFS occurs since the production of cytokines following infection mirror some of the common symptoms of ME/CFS, including central and peripheral fatigue (34). Direct evidence for a role of inflammation in ME/CFS remains limited. A number of studies have examined the potential role of single cytokines or small families of cytokines and a systematic review concluded that attempts to identify individual cytokines as biomarkers of ME/CFS have been inconsistent (7). However, studies do not take into account the interplay of multiple factors, and therefore the diagnostic potential of cytokine profiles has been inconclusive. The cytokine network is a complex, dynamic system and we hypothesised that patterns of cytokine covariation would provide novel insights into the pathophysiology of ME/CFS-related phenotypes. We have employed advanced multivariate analyses to cytokine profiles that leverages covariation and intrinsic redundancy to identify patterns of immune signaling that can be evaluated for their predictions of disease phenotype. The current study identified discriminatory cytokine profiles that can be sufficiently used to distinguish HCs from patients with ME/CFS and provides compelling evidence that a limited number of cytokines can predict with diagnosis and fatigue in these patients. Moreover, this study demonstrates significant potential of using multiplex cytokine profiles and bioinformatics as diagnostic tools for ME/CFS. Thus, particular plasma cytokines, when adjusted for age, gender and cytokine interactions could predict both diagnosis and a number of measures common to patients with ME/CFS including MVC and perceived fatigue as well as cognitive indices such as pattern and verbal reaction times. Specifically, high levels of IL-10, MIP-1β, and RANTES and low levels of PDGF, IL-6, eotaxin, MIP-1α and VEGF were associated with an increased probability of having ME/CFS. With respect to the specific indices of ME/CFS, fatigue and general health, higher plasma values of IL-1β, IL-8, IL-10, IP-10, and RANTES were significantly associated with higher Chalder Fatigue scores i.e. greater fatigue. Higher plasma values of PDGF, eotaxin, G-CSF, and MIP-1α were associated with less fatigue and lower probability of a diagnosis of ME/CFS. Similarly, Higher values of IL-10 and RANTES and lower values of PDGF and MIP-1α were associated with lower SF36 scores (worse physical health). The analysis indicated that higher values of IL-8, IP-10, and TNF-α and lower values of PDGF, IL-1β, eotaxin, and G-CSF were associated with lower MVC. Individuals with ME/CFS were slower in response times and the speed of response was related to MVC, Chalder fatigue and SF36 scores demonstrating a significant interaction between response times and measures of muscle function, perceived fatigue and general health. For example. patients with ME/CFS were slower on MOT, and those who were slower also had poorer MVC, worse fatigue and poorer SF36 physical scores, irrespective of age and gender. Plasma cytokine levels predicted performance on some tests. For example, for VRM-immediate, performance differences were explained by IP-10, IL-7, MIP-1α, IL-13, VEGF and IL-17 levels. Further analyses revealed strong correlations between plasma RANTES and eotaxin levels and the poorer verbal recall and RTs of patients with ME/CFS. Thus, RANTES and eotaxin predict VRM recall. IP-10 was a significant predictor in the presence of 7 significant interactions from 12 possible.

A series of logistic regressions (adjusted for age and gender) confirmed that specific cytokines were able to predict whether an individual was likely to have ME/CFS or be a HC. Age was a significant predictor with older participants classified as more likely to have ME/CFS but gender did not consistently predict likelihood of ME/CFS. In an exploratory cluster analysis, IP-10 and eotaxin were linked whilst RANTES was a separate cluster (see Dendogram, *SI Appendix* Figure S4). In addition to the optimal model for prediction of ME/CFS, we investigated two-way interactions of plasma cytokines in prediction of ME/CFS. We identified 12 two-way interactions that were significant. Among these 12 interactions, six involve IL-6.

The source of plasma cytokines in patients with ME/CFS are unclear, but a number of cytokines can be produced by muscle (75-78). Studies have shown that the acute release of cytokines by muscle during exercise, a process which is quickly resolved, is likely to contribute to the positive effects of activity (78). In contrast, a chronic release of pro-inflammatory cytokines will undoubtedly have a detrimental effect on both muscle and non-muscle organs including the brain, and such cross-tissue interactions have been relatively well described during ageing (77). The specific increased production of IP-10 and MCP-1 by muscle is likely to have an effect on local tissue such as the peripheral nerve. This is supported by studies in inflammatory demyelinating neuropathies which imply a pathogenic role for IP-10 in the genesis of these neuropathies (79). The current study demonstrated no intrinsic deficit in the muscle contractile proteins but demonstrated a deficit in MVC in patients with ME/CFS compared with maximum contractile forces generated by electrical stimulation of lower limb muscles. Failure of voluntary muscle contraction can occur at all levels of the neuromuscular system, but it is tempting to speculate that the local increased production of specific cytokines may modulate the function of motoneuron. The identification of an increase in the number of small muscle fibres in muscles of patients with ME/CFS would also be consistent with active denervation of a small number of muscle fibres (80) although this was not studied in detail.

The nature of the discriminatory cytokines in this study is intriguing. There is growing evidence of the prominent role of low-grade chronic inflammation in age-related changes in the neuromuscular system (81). The inflammatory immune response and cytokine levels have been associated with both fatigue and depression for some time and studies have suggested that changes in plasma levels of a range of cytokines in certain conditions may be predictive of future depression (14). The consistent association of higher levels of RANTES, IP-10 and IL-10 and lower levels of eotaxin, MIP1α and PDGF with a number of physiological and cognitive features of ME/CFS provides compelling evidence for a role of these cytokine/chemokines in the pathology of ME/CFS. IL-10 is a cytokine with potent anti-inflammatory properties produced by a wide range of cells that plays a central role in limiting host immune response to pathogens, preventing damage to the host and maintaining normal tissue homeostasis. Dysregulation of IL-10 is associated with enhanced immunopathology in response to infection as well as increased risk for development of many autoimmune diseases (82). The association of an elevated level of IL-10 with diagnosis and physiological features of ME/CFS are counterintuitive, but some pathogens can harness the immunosuppressive capacity of IL-10 to limit host immune response, leading to persistent infection (82). RANTES, MIP-1α and MIP-1β are inflammatory cytokines, co-secreted *de novo* by immune cells with infection (83) and several studies of the levels of these three cytokines in patients with ME/CSF show that they are all changed in the same direction together (7). Both RANTES and MIP1β were predictive of diagnosis of ME/CFS in the model used in the current study. In a much smaller ME/CFS cohort study, serum RANTES was elevated in patients with ‘moderate’ ME/CFS (84). Studies examining the effect of RANTES on skeletal muscle function are scarce but Kohno *et al* (85) demonstrated an inhibitory effect of CD8-positive T cell production of RANTES on muscle regeneration. Compelling evidence for a role of IP-10 in the pathology of CFS comes from a landmark study by Hornig et al (86) which examined the cerebral spinal fluid (CSF) of patients with ME/CFS in comparison with patients with multiple sclerosis (MS) and HCs. The CSF of patients with ME/CFS demonstrated a pattern of changes consistent with immune activation in the CNS. Marked changes were seen in IP-10 levels, specifically, IP-10 levels in ME/CFS were elevated to a similar degree to those seen in MS. There are multiple reports that circulating IP-10 levels are increased in humans and mice infected with either viruses (87) or bacteria (88). Increases in IP-10 are found in patients with Behcet’s syndrome (89), hepatitis C (90), tuberculosis (TB) (91) and Leishmaniosis (92). IP-10 has also been implicated in inflammatory diseases (93) and in numerous autoimmune diseases (94). Blood IP-10 concentrations have been proposed as a biomarker of disease activity that predicts response to treatment for TB, HIV and hepatitis C, where patients with lower IP-10 showed a more rapid virologic response and intracellular infections are controlled better or cleared faster in the absence of IL-10 (95). Other researchers have shown that treatment response to hepatitis B and C is independently associated with IP-10 polymorphisms (96, 97). In muscle cells infected with different viruses, IP-10 was among the most highly upregulated genes (98). Furthermore, an intervention to decrease IP-10 was found to reduce inflammation in a mouse model of myositis (99). The association of low levels of eotaxin and PDGF with physiological and cognitive features of ME/CFS is difficult to interpret. Evidence indicates that increased levels of eotaxin-1 and IP-10 are associated with euthymia (100). Platelet-derived growth-factor (PDGF) has been shown to promote blood vessel growth and neuronal survival (101, 102). Eotaxin is also by a wide range of cells, including immune and cells of the CNS and is a potent eosinophil chemoattractant cytokine. PDGF, together with Insulin growth factor I (IGF-I) play an important role in muscle regeneration (78). The level of circulating PDGF and eotaxin is lower in the elderly (103, 104) although there are no studies examining the direct effect of this reduced level. The mechanisms by which changes in circulating levels of pro-inflammatory cytokines exert their effects on cognition are unclear, but it has been proposed that cytokines may induce some changes at the blood brain barrier. Studies in mice have shown that TNFα causes changes in the endothelial cells constituting the blood brain barrier, resulting in increased permeability in animal models, an effect which can be restored by treatment with anti-inflammatory drugs (105).

In summary, this original and extensive study demonstrated that although there was little dysfunction evident in skeletal muscle mitochondria of patients with ME/CFS, particular cytokines, when adjusted for age, gender and cytokine interactions could predict both diagnosis and a number of measures common to patients with ME/CFS. These included MVC and perceived fatigue as well as cognitive indices such as pattern and verbal reaction times. Further, data demonstrated that muscle was a major source of IP-10 and MCP-1. We employed advanced multivariate analyses to cytokine profiles that leverages covariation and intrinsic redundancy to identify patterns of immune signaling that can be evaluated for their predictions of disease phenotype. The current study identified discriminatory cytokine profiles that can be sufficiently used to distinguish HCs from patients with ME/CFS and provides compelling evidence that a limited number of cytokines are associated with diagnosis and fatigue. Moreover, this study demonstrates significant potential of using multiplex cytokine profiles and bioinformatics as diagnostic tools for ME/CFS. Multi-biomarker-based diagnostics tests, including those using cytokines are impacting patient care and outcomes in a number of illnesses (106), potentiating the possibility of use of this model not only in diagnosis, but also being able to individually personalise therapies. With the potential of increasing numbers of patients developing ME/CFS during or following Coronavirus disease 2019 (COVID-19), caused by SARS-CoV-2, further studies will be vital to assess the similarities in pathophysiology with those described here.

## Data Availability

The datasets generated during and/or analysed during the current study are available from the corresponding author on reasonable request.

## Acknowledgements

The authors would like to thank the MRC for their generous financial support of this work. The work would not be possible without the dedicated work of the ME/CFS Diagnostic and Therapy Team, including Collette Foster, Claire Moss, Debbie Gardner, Catherine Wallace, Deb Robert, Jen Pomfrett, Jayne Galvin, Jo Dent, Jane Knight, Jane Hillard and Jeannie Battard.

---

**Supplementary Figure S1.**
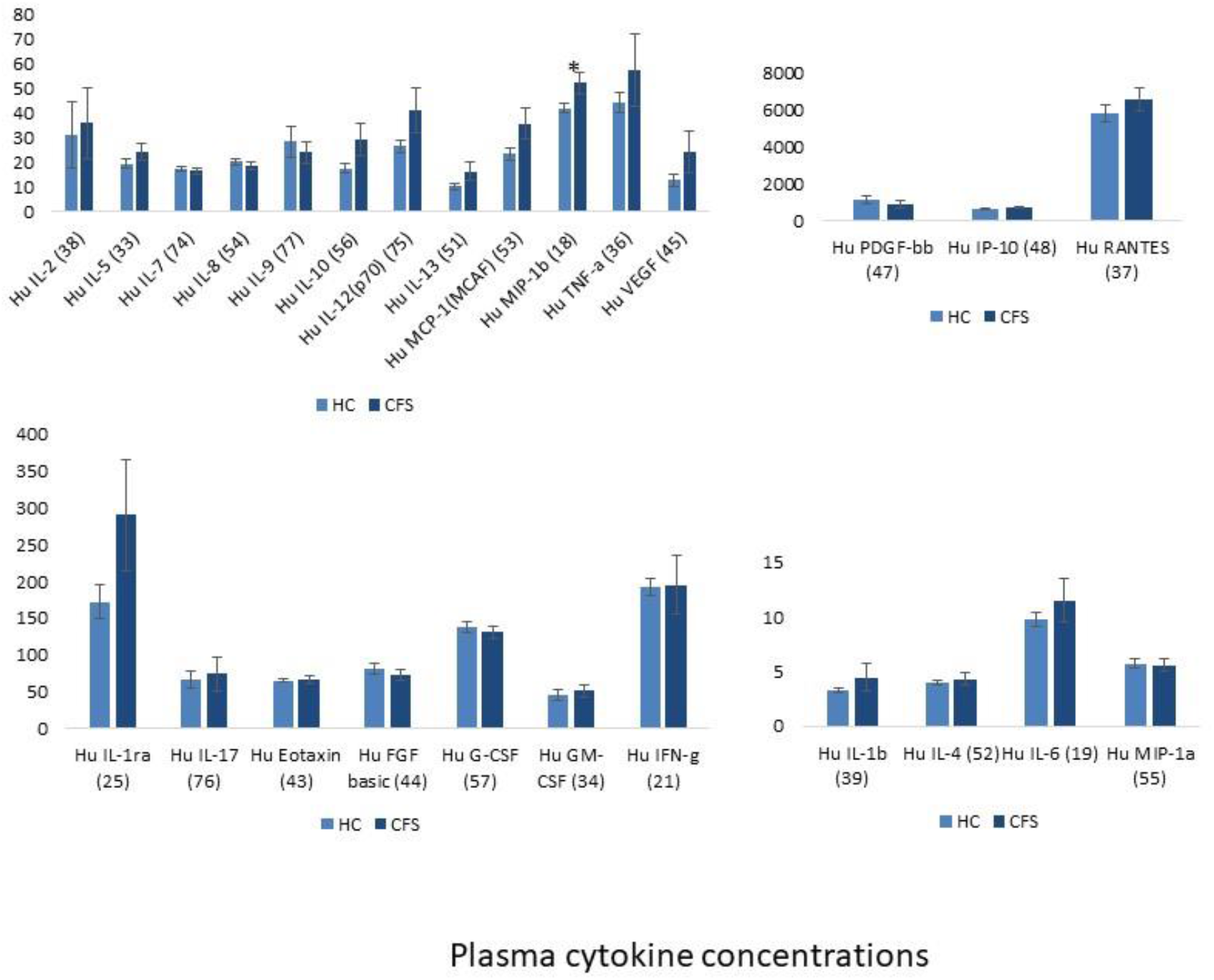
Individual cytokine concentrations in plasma of patients with ME/CFS and HCs. Data presented as mean ± SEM. The asterisks (*) indicate p-values less than 0.05.

**Supplementary Figure S2.**
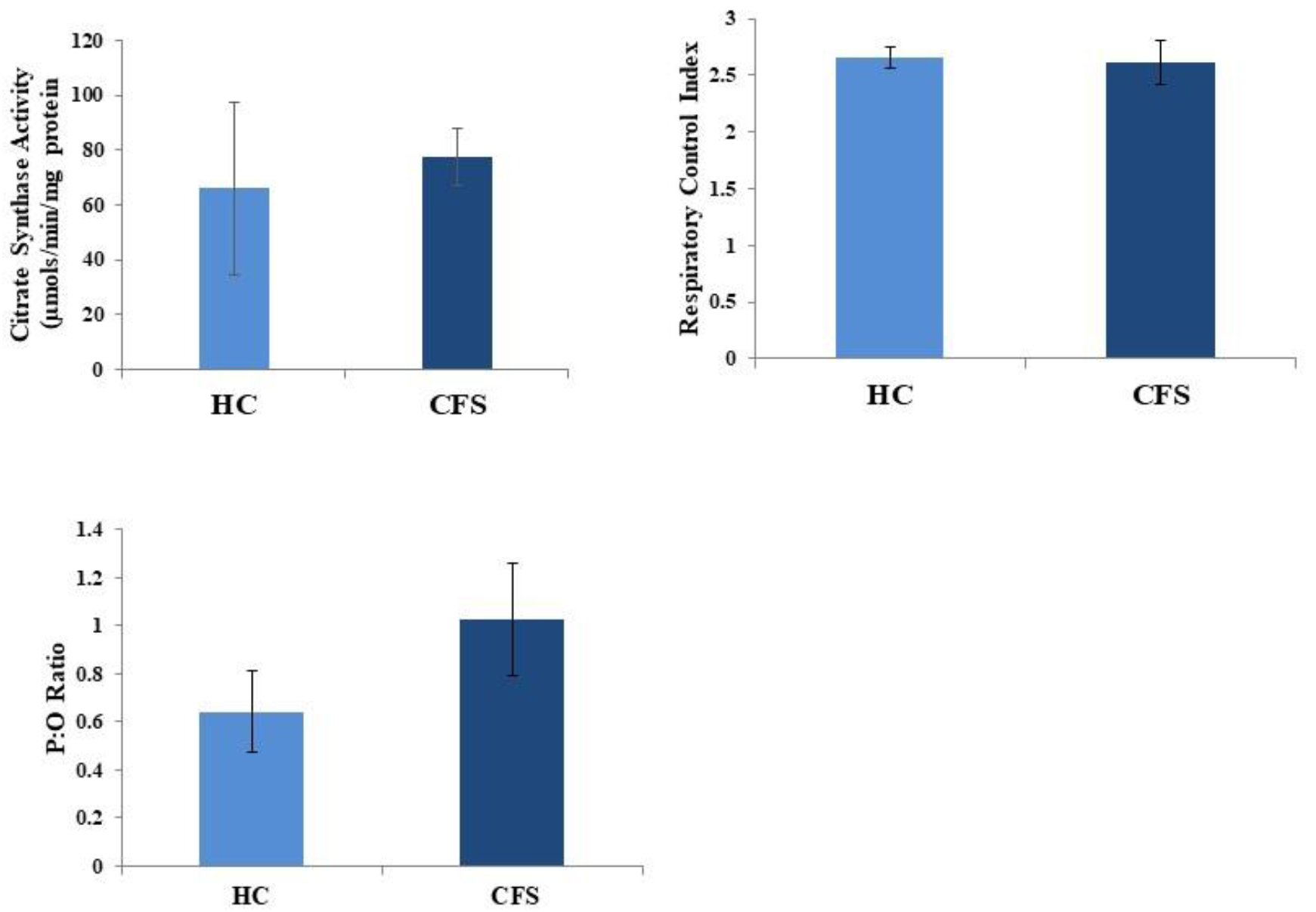
**(A)** Citrate synthase activity of muscle homogenates from *vastus lateralis* (VL) from in patients with ME/CFS and HCs. **(B)** Respiratory control index (RCI) of intact mitochondria in permeabilised myofibres prepared from *vastus lateralis* (*VL*) from patients with ME/CFS and HCs. **(C)** P:O Ratio of ATP formed against oxygen utilised of intact mitochondria in permeabilised myofibres prepared from *VL* of patients with ME/CFS and HCs. Data presented as mean ± SEM.

**Supplementary Figure S3.**
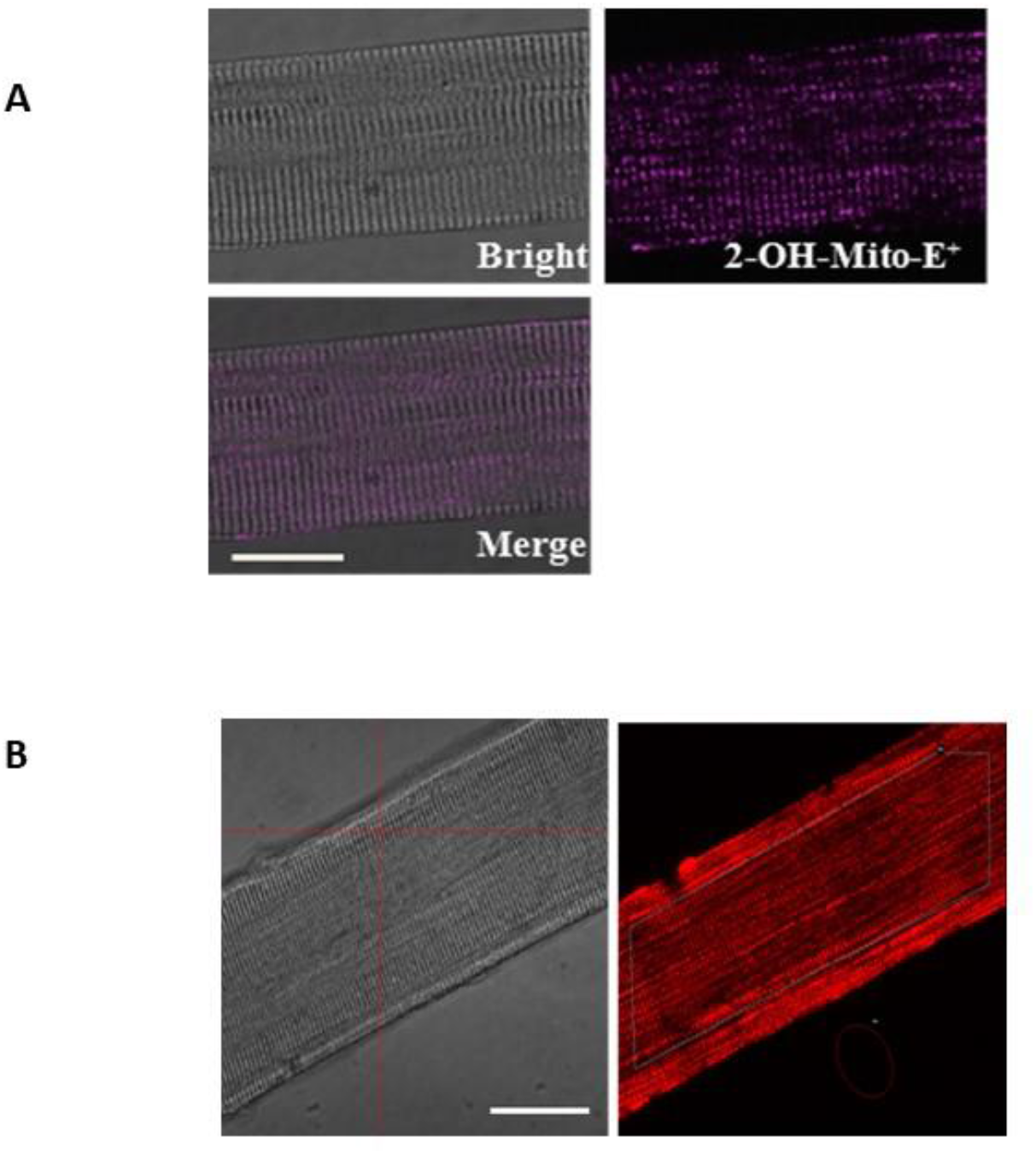
(A) Representative images of a single fibre isolated from the VL muscle under bright field, fluorescent image following loading with MitoSOX Red (20nM, Purple), and a merged image as indicated and analysed by confocal microscopy. 60x original magnification. Scale bar, 25μm. (B) Representative confocal images of an isolated fibre showing TMRM fluorescence. 60x original magnification. Scale bar, 25μm.

**Supplementary Figure S4.**
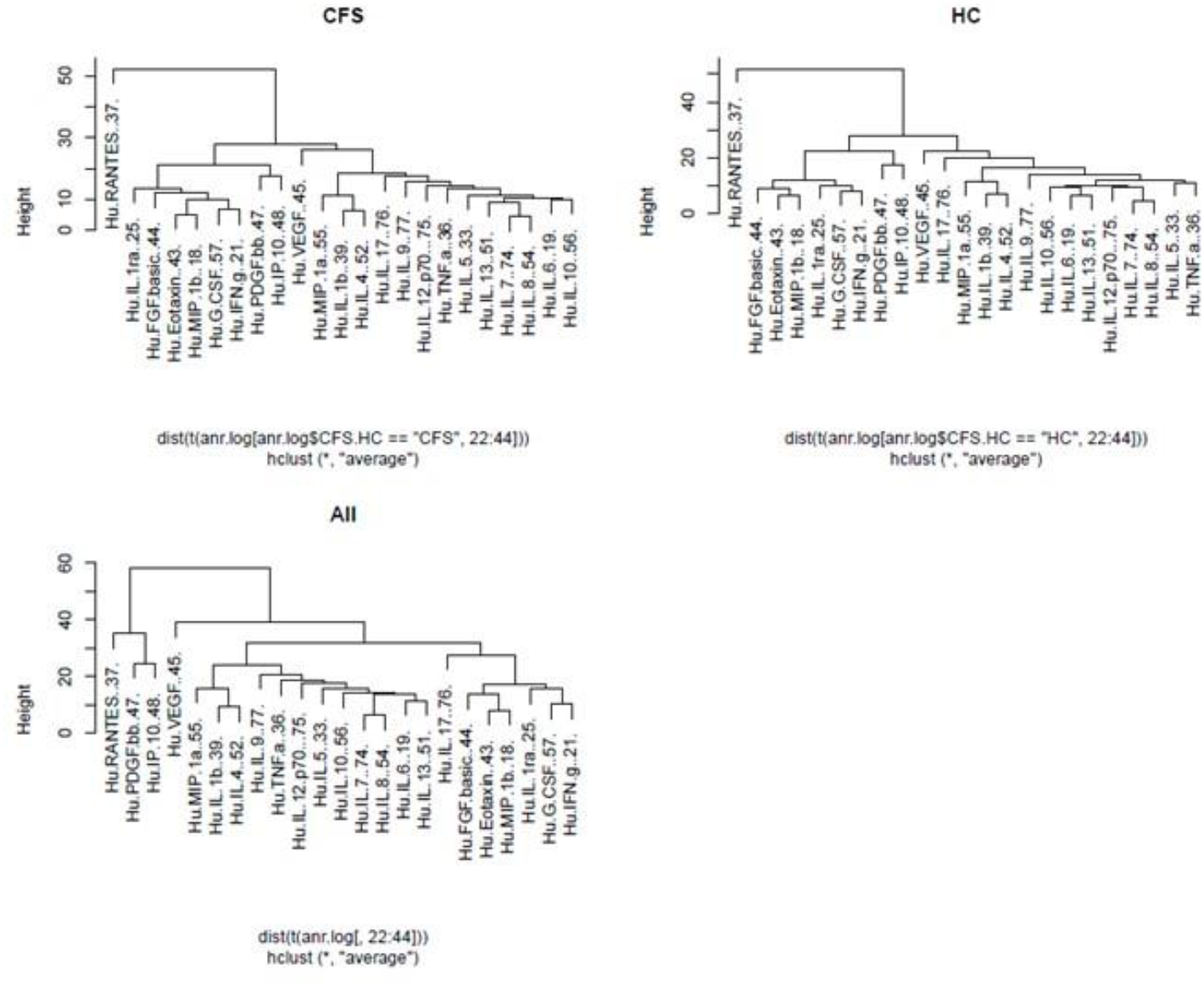
Hierarchical cluster analysis of plasma cytokines, separated by CFS/HC and all.

**Supplementary Table S1.** Inclusion and Exclusion criteria for study participation.

| Inclusion Criteria | Exclusion Criteria |
| --- | --- |
| Patients should be newly diagnosed with CFS and will be diagnosed with CFS to meet the CDC, Oxford Criteria (1990) and recommended NICE guidelines (2007). | Patients with history of substance misuse |
| Age between 18-55 years | Smokers |
| Definitive onset i.e. it is not lifelong | Patients taking antioxidant vitamin supplements |
| Fatigue must be the main symptom | Patients who are non-ambulatory or have pre-existing on-going mental health illness, including levels of anxiety and depression as described by the HADS scale (1983) |
| The fatigue should be severe, disabling and affect both physical and mental functioning i.e. significant impairment of short-term memory or concentration |  |
| The fatigue must be present for at least 6 months, during which it has to have been present for more than 50 % of the time |  |
| Other symptoms may be present: Particularly Myalgia, Sore throat, Tender lymph glands in the neck region or under the armpits, Headaches of a new type, pattern and severity, Malaise following effort that lasts more than 24 hrs, Pains in several joints occurring without swelling or redness, Mood and sleep disturbance, Palpitations without heart disease, Dizziness, with or without nausea. |  |
| FBC and Clotting screen within normal laboratory range |  |
| Able to provide written informed consent. |  |

**Supplementary Table S2.** Symptom assessment of patients with ME/CFS. Data presented as % of study population.

|  | **CFS (%)** |  | **CFS (%)** |
| --- | --- | --- | --- |
| **Sleep disturbances** | 64 | **Post exertional malaise** | 100 |
| **Muscle pain** | 42 | **Cognitive dysfunction** | 29 |
| **Joint pain** | 33 | **General malaise/flu-like** | 29 |
| **Headaches** | 35 | **Dizziness** | 25 |
| **Painful lymph nodes** | 7 | **Nausea** | 17 |
| **Sore Throat** | 17 | **Palpitations** | 8 |

**Supplementary Table S3.**
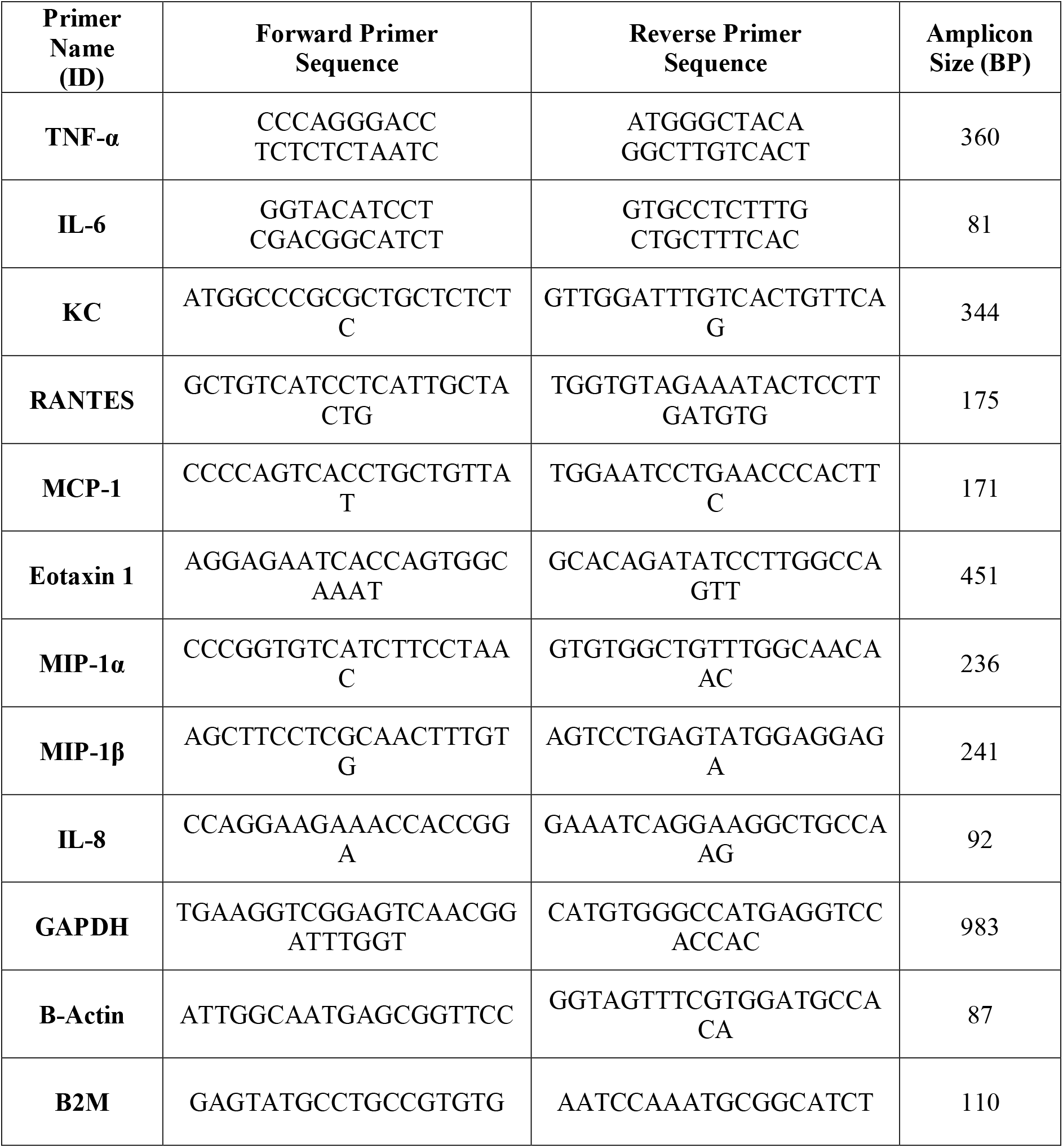
Sequences of the specific primers used for q-PCR amplification of cytokines, atrogin-1 and housekeeping genes.

**SI appendix Table S4.**
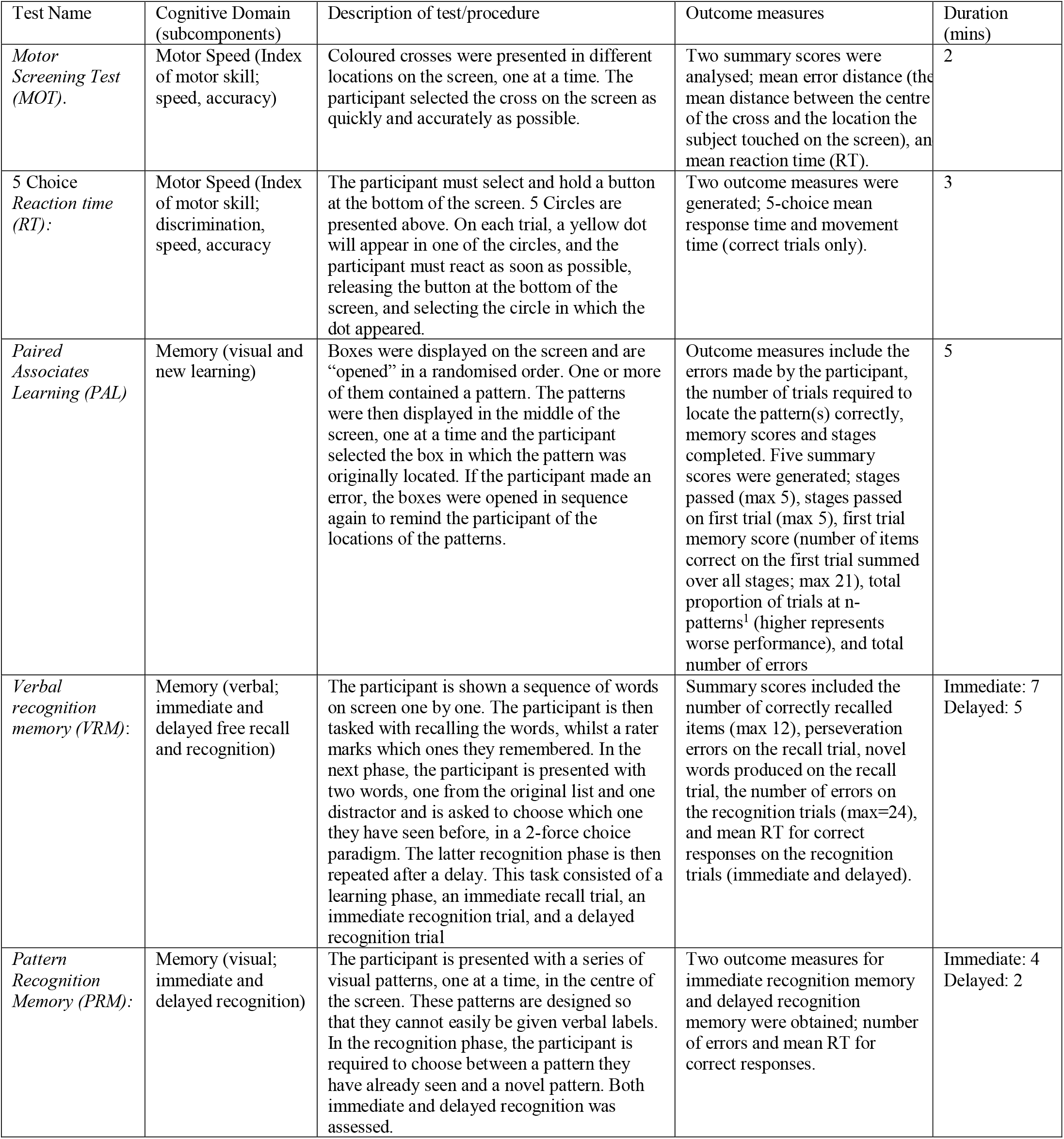
Description of tests administered from the Cambridge Automated Europsychological Test Battery (CANTAB, Cambridge Cognition, UK).

**Supplemental Table S5.**
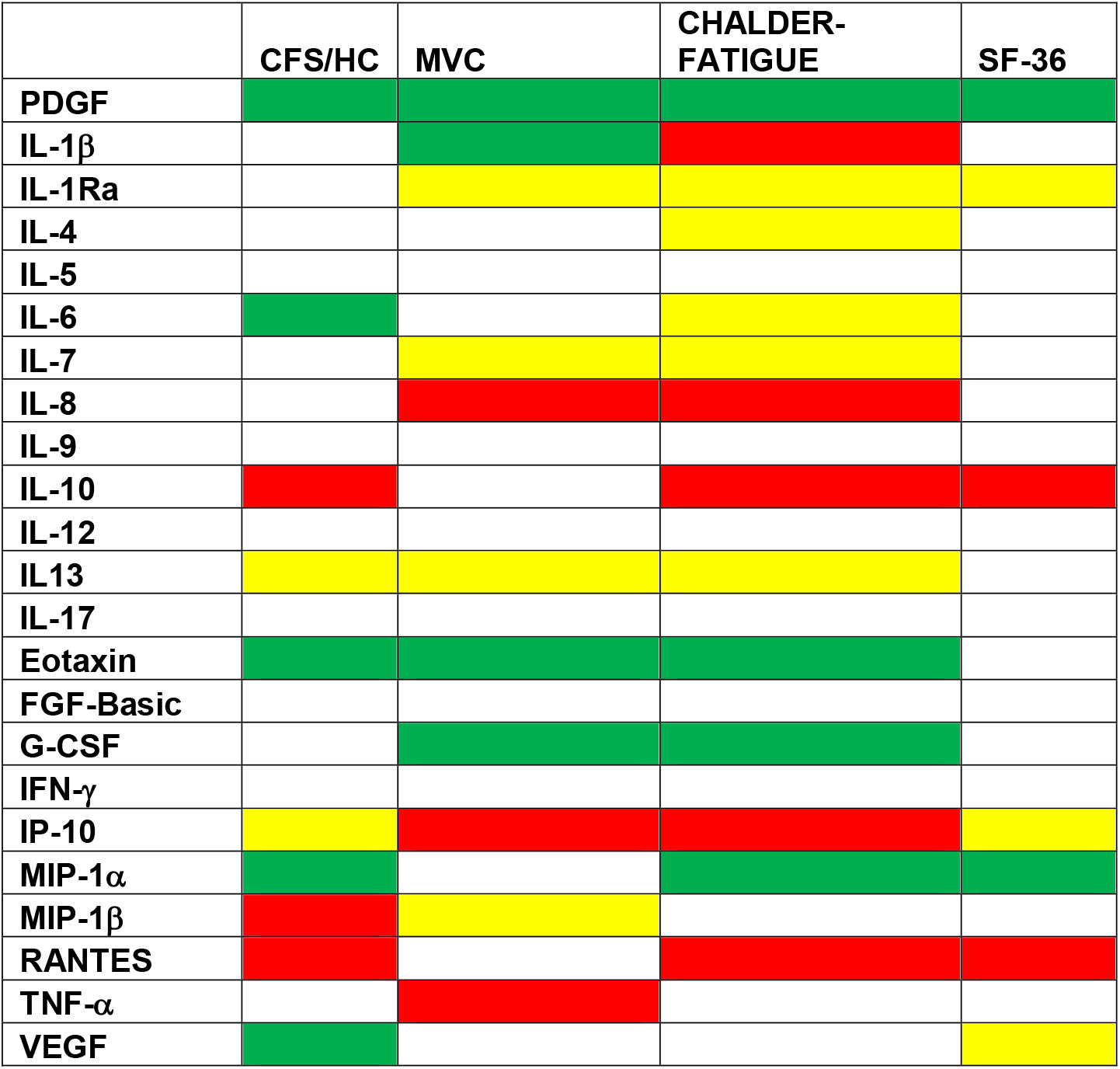
Directional changes in cytokines associated with ME/CFS diagnosis, low MVC, high Chalder Fatigue score (more fatigued) or low SF-36 score (worse health). Red indicates that increasing levels of cytokines are significantly associated with worsening prognosis: increased risk of ME/CFS, more fatigue (increasing Chalder Fatigue score), worse physical health (lowering SF-36 score), and worse muscle function (lower MVC). Green colours indicate that levels of cytokines are significantly associated with better prognosis: lowered risk of ME/CFS, less fatigue (lowering Chalder Fatigue score), better physical health (increasing SF-36 score), and better muscle function (increasing MVC). Yellow colours indicate that the cytokines are in the model but non-significant, blank indicates not in the model.

**Supplementary Table S6.**
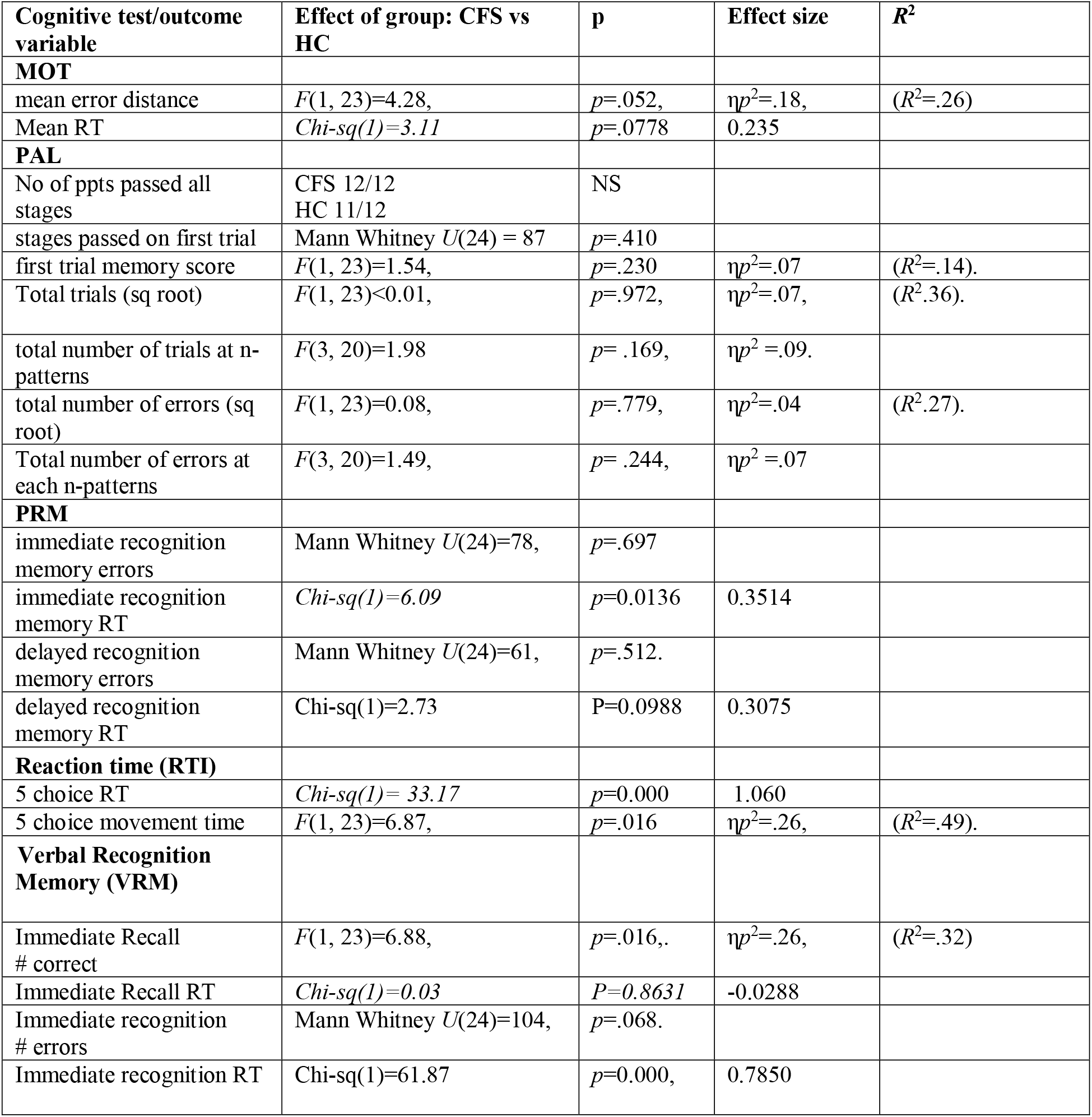
Comparison of ME/CFS vs. HC on all cognitive outcomes from the Cambridge Automated Neuropsychological Test Battery (Cantab, Cambridge Cognition, UK), adjusted for age and sex.

**Supplementary Table S7.**
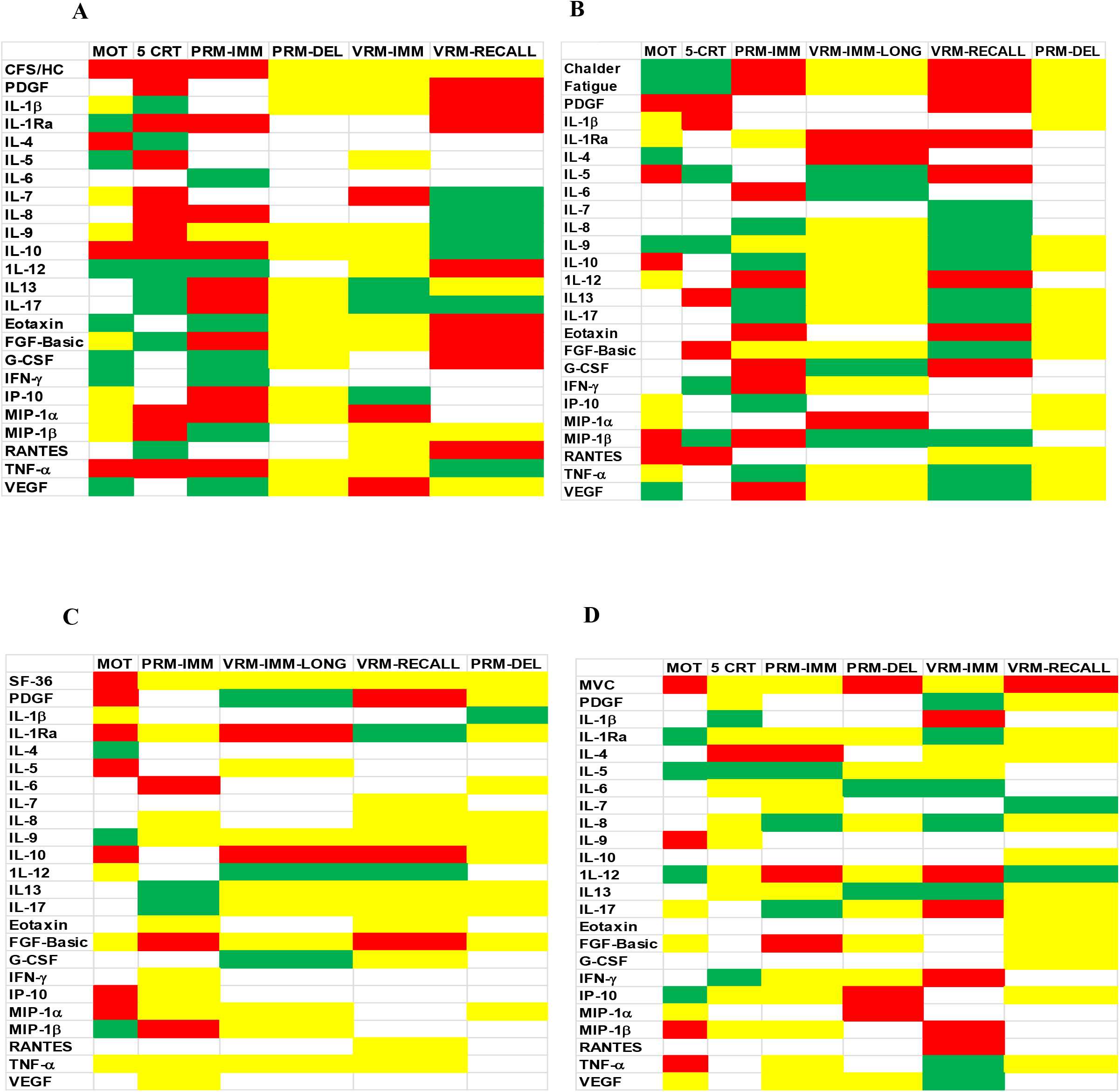
Direction of responses in cytokines associated with cognitive tests.

## Footnotes

1 Total proportion of trials was used as there were a different number of problems at each length - 2 problems with 2 patterns but only one with 3-8 patterns.

## References

1. NICE. Chronic fatigue syndrome/myalgic encephalomyelitis (or encephalopathy): Diagnosis and management of CFS/ME in adults and children. 2010 [Available from: http://guidanceniceorguk/CG53/NICEGuidance/pdf/English.

2. Sandler CX, Lloyd AR. Chronic fatigue syndrome: progress and possibilities. Med J Aust. 2020.

3. Bested AC, Marshall LM. Review of Myalgic Encephalomyelitis/Chronic Fatigue Syndrome: an evidence-based approach to diagnosis and management by clinicians. Rev Environ Health. 2015;30(4):223–49.

4. Jason LA, Richman JA, Rademaker AW, Jordan KM, Plioplys AV, Taylor RR, et al. A community-based study of chronic fatigue syndrome. Arch Intern Med. 1999;159(18):2129–37.

5. Nacul LC, Lacerda EM, Pheby D, Campion P, Molokhia M, Fayyaz S, et al. Prevalence of myalgic encephalomyelitis/chronic fatigue syndrome (ME/CFS) in three regions of England: a repeated cross-sectional study in primary care. BMC Med. 2011;9:91.

6. Johnston S, Brenu EW, Staines DR, Marshall-Gradisnik S. The adoption of chronic fatigue syndrome/myalgic encephalomyelitis case definitions to assess prevalence: a systematic review. Ann Epidemiol. 2013;23(6):371–6.

7. Blundell S, Ray KK, Buckland M, White PD. Chronic fatigue syndrome and circulating cytokines: A systematic review. Brain Behav Immun. 2015;50:186-95.

8. Blomberg J, Gottfries CG, Elfaitouri A, Rizwan M, Rosen A. Infection Elicited Autoimmunity and Myalgic Encephalomyelitis/Chronic Fatigue Syndrome: An Explanatory Model. Front Immunol. 2018;9:229.

9. Magnus P, Gunnes N, Tveito K, Bakken IJ, Ghaderi S, Stoltenberg C, et al. Chronic fatigue syndrome/myalgic encephalomyelitis (CFS/ME) is associated with pandemic influenza infection, but not with an adjuvanted pandemic influenza vaccine. Vaccine. 2015;33(46):6173–7.

10. Underhill RA. Myalgic encephalomyelitis, chronic fatigue syndrome: An infectious disease. Med Hypotheses. 2015;85(6):765–73.

11. Hickie I, Davenport T, Wakefield D, Vollmer-Conna U, Cameron B, Vernon SD, et al. Post-infective and chronic fatigue syndromes precipitated by viral and non-viral pathogens: prospective cohort study. Bmj. 2006;333(7568):575.

12. Tsai SY, Yang TY, Chen HJ, Chen CS, Lin WM, Shen WC, et al. Increased risk of chronic fatigue syndrome following herpes zoster: a population-based study. Eur J Clin Microbiol Infect Dis. 2014;33(9):1653–9.

13. Brurberg KG, Fonhus MS, Larun L, Flottorp S, Malterud K. Case definitions for chronic fatigue syndrome/myalgic encephalomyelitis (CFS/ME): a systematic review. BMJ Open. 2014;4(2):e003973.

14. Lee CH, Giuliani F. The Role of Inflammation in Depression and Fatigue. Front Immunol. 2019;10:1696.

15. Morris G, Maes M, Berk M, Puri BK. Myalgic encephalomyelitis or chronic fatigue syndrome: how could the illness develop? Metab Brain Dis. 2019;34(2):385–415.

16. Picca A, Mankowski RT, Burman JL, Donisi L, Kim JS, Marzetti E, et al. Mitochondrial quality control mechanisms as molecular targets in cardiac ageing. Nat Rev Cardiol. 2018;15(9):543–54.

17. Earl KE, Sakellariou GK, Sinclair M, Fenech M, Croden F, Owens DJ, et al. Vitamin D status in chronic fatigue syndrome/myalgic encephalomyelitis: a cohort study from the North-West of England. BMJ Open. 2017;7(11):e015296.

18. Pye D, Palomero J, Kabayo T, Jackson MJ. Real-time measurement of nitric oxide in single mature mouse skeletal muscle fibres during contractions. J Physiol. 2007;581(Pt 1):309-18.

19. Palomero J, Pye D, Kabayo T, Spiller DG, Jackson MJ. In situ detection and measurement of intracellular reactive oxygen species in single isolated mature skeletal muscle fibers by real time fluorescence microscopy. Antioxid Redox Signal. 2008;10(8):1463–74.

20. Powers SK, Jackson MJ. Exercise-induced oxidative stress: cellular mechanisms and impact on muscle force production. Physiol Rev. 2008;88(4):1243–76.

21. Debold EP. Potential molecular mechanisms underlying muscle fatigue mediated by reactive oxygen and nitrogen species. Front Physiol. 2015;6:239.

22. Kuwahara H, Horie T, Ishikawa S, Tsuda C, Kawakami S, Noda Y, et al. Oxidative stress in skeletal muscle causes severe disturbance of exercise activity without muscle atrophy. Free Radic Biol Med. 2010;48(9):1252–62.

23. Lustgarten MS, Jang YC, Liu Y, Qi W, Qin Y, Dahia PL, et al. MnSOD deficiency results in elevated oxidative stress and decreased mitochondrial function but does not lead to muscle atrophy during aging. Aging Cell. 2011;10(3):493–505.

24. Kramer P, Bressan P. Our (Mother’s) Mitochondria and Our Mind. Perspect Psychol Sci. 2018;13(1):88–100.

25. Tarantini S, Valcarcel-Ares NM, Yabluchanskiy A, Fulop GA, Hertelendy P, Gautam T, et al. Treatment with the mitochondrial-targeted antioxidant peptide SS-31 rescues neurovascular coupling responses and cerebrovascular endothelial function and improves cognition in aged mice. Aging Cell. 2018;17(2).

26. Lomeli N, Di K, Czerniawski J, Guzowski JF, Bota DA. Cisplatin-induced mitochondrial dysfunction is associated with impaired cognitive function in rats. Free Radic Biol Med. 2017;102:274-86.

27. Behan WM, More IA, Behan PO. Mitochondrial abnormalities in the postviral fatigue syndrome. Acta Neuropathol. 1991;83(1):61–5.

28. Myhill S, Booth NE, McLaren-Howard J. Chronic fatigue syndrome and mitochondrial dysfunction. Int J Clin Exp Med. 2009;2(1):1–16.

29. Plioplys AV, Plioplys S. Electron-microscopic investigation of muscle mitochondria in chronic fatigue syndrome. Neuropsychobiology. 1995;32(4):175–81.

30. Barnes PR, Taylor DJ, Kemp GJ, Radda GK. Skeletal muscle bioenergetics in the chronic fatigue syndrome. J Neurol Neurosurg Psychiatry. 1993;56(6):679–83.

31. Kaplan CM, Schrepf A, Ichesco E, Larkin T, Harte SE, Harris RE, et al. Inflammation is associated with pro-nociceptive brain connections in rheumatoid arthritis patients with concomitant fibromyalgia. Arthritis Rheumatol. 2019.

32. Missailidis D, Annesley SJ, Fisher PR. Pathological Mechanisms Underlying Myalgic Encephalomyelitis/Chronic Fatigue Syndrome. Diagnostics (Basel). 2019;9(3).

33. Skougaard M, Jorgensen TS, Rifbjerg-Madsen S, Coates LC, Egeberg A, Amris K, et al. In Psoriatic Arthritis fatigue is driven by inflammation, disease duration, and chronic pain: An observational DANBIO registry study. J Rheumatol. 2019.

34. VanElzakker MB, Brumfield SA, Lara Mejia PS. Neuroinflammation and Cytokines in Myalgic Encephalomyelitis/Chronic Fatigue Syndrome (ME/CFS): A Critical Review of Research Methods. Front Neurol. 2018;9:1033.

35. Maes M, Mihaylova I, Kubera M, Bosmans E. Not in the mind but in the cell: increased production of cyclo-oxygenase-2 and inducible NO synthase in chronic fatigue syndrome. Neuro Endocrinol Lett. 2007;28(4):463–9.

36. Broderick G, Fuite J, Kreitz A, Vernon SD, Klimas N, Fletcher MA. A formal analysis of cytokine networks in chronic fatigue syndrome. Brain Behav Immun. 2010;24(7):1209–17.

37. Lorusso L, Mikhaylova SV, Capelli E, Ferrari D, Ngonga GK, Ricevuti G. Immunological aspects of chronic fatigue syndrome. Autoimmun Rev. 2009;8(4):287–91.

38. Montoya JG, Holmes TH, Anderson JN, Maecker HT, Rosenberg-Hasson Y, Valencia IJ, et al. Cytokine signature associated with disease severity in chronic fatigue syndrome patients. Proc Natl Acad Sci U S A. 2017;114(34):E7150-e8.

39. Ghosh N, Ghosh R, Mandal SC. Antioxidant protection: A promising therapeutic intervention in neurodegenerative disease. Free Radic Res. 2011;45(8):888–905.

40. Reid MB, Moylan JS. Beyond atrophy: redox mechanisms of muscle dysfunction in chronic inflammatory disease. J Physiol. 2011;589(Pt 9):2171-9.

41. Villeda SA, Luo J, Mosher KI, Zou B, Britschgi M, Bieri G, et al. The ageing systemic milieu negatively regulates neurogenesis and cognitive function. Nature. 2011;477(7362):90–4.

42. Castellano JM, Kirby ED, Wyss-Coray T. Blood-Borne Revitalization of the Aged Brain. JAMA Neurol. 2015;72(10):1191–4.

43. de Miranda AS, Brant F, Campos AC, Vieira LB, Rocha NP, Cisalpino D, et al. Evidence for the contribution of adult neurogenesis and hippocampal cell death in experimental cerebral malaria cognitive outcome. Neuroscience. 2015;284:920-33.

44. Lightfoot AP, Sakellariou GK, Nye GA, McArdle F, Jackson MJ, Griffiths RD, et al. SS-31 attenuates TNF-alpha induced cytokine release from C2C12 myotubes. Redox Biol. 2015;6:253-9.

45. Sharpe MC, Archard LC, Banatvala JE, Borysiewicz LK, Clare AW, David A, et al. A report-chronic fatigue syndrome: guidelines for research. J R Soc Med. 1991;84(2):118–21.

46. Fukuda K, Straus SE, Hickie I, Sharpe MC, Dobbins JG, Komaroff A. The chronic fatigue syndrome: a comprehensive approach to its definition and study. International Chronic Fatigue Syndrome Study Group. Ann Intern Med. 1994;121(12):953–9.

47. Drouin JM, Valovich-mcLeod TC, Shultz SJ, Gansneder BM, Perrin DH. Reliability and validity of the Biodex system 3 pro isokinetic dynamometer velocity, torque and position measurements. Eur J Appl Physiol. 2004;91(1):22–9.

48. Trappe S, Gallagher P, Harber M, Carrithers J, Fluckey J, Trappe T. Single muscle fibre contractile properties in young and old men and women. J Physiol. 2003;552(Pt 1):47-58.

49. Sakellariou GK, Pearson T, Lightfoot AP, Nye GA, Wells N, Giakoumaki, II, et al. Long-term administration of the mitochondria-targeted antioxidant mitoquinone mesylate fails to attenuate age-related oxidative damage or rescue the loss of muscle mass and function associated with aging of skeletal muscle. Faseb j. 2016;30(11):3771–85.

50. Gouspillou G, Sgarioto N, Kapchinsky S, Purves-Smith F, Norris B, Pion CH, et al. Increased sensitivity to mitochondrial permeability transition and myonuclear translocation of endonuclease G in atrophied muscle of physically active older humans. Faseb j. 2014;28(4):1621–33.

51. Degens H, Bosutti A, Gilliver SF, Slevin M, van Heijst A, Wust RC. Changes in contractile properties of skinned single rat soleus and diaphragm fibres after chronic hypoxia. Pflugers Arch. 2010;460(5):863–73.

52. Lynch GS, Faulkner JA, Brooks SV. Force deficits and breakage rates after single lengthening contractions of single fast fibers from unconditioned and conditioned muscles of young and old rats. Am J Physiol Cell Physiol. 2008;295(1):C249-56.

53. Gnaiger E, Mendez G, Hand SC. High phosphorylation efficiency and depression of uncoupled respiration in mitochondria under hypoxia. Proc Natl Acad Sci U S A. 2000;97(20):11080–5.

54. Anderson EJ, Neufer PD. Type II skeletal myofibers possess unique properties that potentiate mitochondrial H(2)O(2) generation. Am J Physiol Cell Physiol. 2006;290(3):C844-51.

55. Vasilaki A, Mansouri A, Van Remmen H, van der Meulen JH, Larkin L, Richardson AG, et al. Free radical generation by skeletal muscle of adult and old mice: effect of contractile activity. Aging Cell. 2006;5(2):109–17.

56. Pearson T, Kabayo T, Ng R, Chamberlain J, McArdle A, Jackson MJ. Skeletal muscle contractions induce acute changes in cytosolic superoxide, but slower responses in mitochondrial superoxide and cellular hydrogen peroxide. PLoS One. 2014;9(5):e96378.

57. Pfeiffer S, Leopold E, Schmidt K, Brunner F, Mayer B. Inhibition of nitric oxide synthesis by NG-nitro-L-arginine methyl ester (L-NAME): requirement for bioactivation to the free acid, NG-nitro-L-arginine. Br J Pharmacol. 1996;118(6):1433–40.

58. Livak KJ, Schmittgen TD. Analysis of relative gene expression data using real-time quantitative PCR and the 2(-Delta Delta C(T)) Method. Methods. 2001;25(4):402–8.

59. Vasilaki A, Simpson D, McArdle F, McLean L, Beynon RJ, Van Remmen H, et al. Formation of 3-nitrotyrosines in carbonic anhydrase III is a sensitive marker of oxidative stress in skeletal muscle. Proteomics Clin Appl. 2007;1(4):362–72.

60. Sakellariou GK, McDonagh B, Porter H, Giakoumaki, II, Earl KE, Nye GA, et al. Comparison of Whole Body SOD1 Knockout with Muscle-Specific SOD1 Knockout Mice Reveals a Role for Nerve Redox Signaling in Regulation of Degenerative Pathways in Skeletal Muscle. Antioxid Redox Signal. 2018;28(4):275–95.

61. Team RC. R: A Language and Environment for Statistical Computing Vienna, Austria: R Foundation for Statistical Computing; 2020 [

62. Pawitan Y. In All Likelihood: Statistical Modelling and Inference Using Likelihood. New York, USA: Oxford University Press:; 2013.

63. Cox D. Regression Models and Life-Tables. Journal of the Royal Statistical Society Series B (Methodological). 1972; 34(2):187-220.

64. T T. A Package for Survival Analysis in R. R package version 3.1-12. 2020.

65. Colett D. Modelling Survival Data in Medical Research, Third Edition.. Boca Raton, USA: Chapman & Hall; 2014.

66. Reyes M, Nisenbaum R, Hoaglin DC, Unger ER, Emmons C, Randall B, et al. Prevalence and incidence of chronic fatigue syndrome in Wichita, Kansas. Arch Intern Med. 2003;163(13):1530–6.

67. Schillings ML, Kalkman JS, van der Werf SP, van Engelen BG, Bleijenberg G, Zwarts MJ. Diminished central activation during maximal voluntary contraction in chronic fatigue syndrome. Clin Neurophysiol. 2004;115(11):2518–24.

68. Light AR, Vierck CJ, Light KC. Frontiers in Neuroscience Myalgia and Fatigue: Translation from Mouse Sensory Neurons to Fibromyalgia and Chronic Fatigue Syndromes. In: Kruger L, Light AR, editors. Translational Pain Research: From Mouse to Man. Boca Raton, FL: CRC Press/Taylor & Francis Llc.; 2010.

69. Gumucio JP, Mendias CL. Atrogin-1, MuRF-1, and sarcopenia. Endocrine. 2013;43(1):12–21.

70. Picard M, Taivassalo T, Ritchie D, Wright KJ, Thomas MM, Romestaing C, et al. Mitochondrial structure and function are disrupted by standard isolation methods. PLoS One. 2011;6(3):e18317.

71. Saks VA, Veksler VI, Kuznetsov AV, Kay L, Sikk P, Tiivel T, et al. Permeabilized cell and skinned fiber techniques in studies of mitochondrial function in vivo. Mol Cell Biochem. 1998;184(1-2):81-100.

72. Geto Z, Molla MD, Challa F, Belay Y, Getahun T. Mitochondrial Dynamic Dysfunction as a Main Triggering Factor for Inflammation Associated Chronic Non-Communicable Diseases. J Inflamm Res. 2020;13:97-107.

73. Piantadosi CA. Mitochondrial DNA, oxidants, and innate immunity. Free Radic Biol Med. 2020;152:455-61.

74. Teodoro T, Edwards MJ, Isaacs JD. A unifying theory for cognitive abnormalities in functional neurological disorders, fibromyalgia and chronic fatigue syndrome: systematic review. J Neurol Neurosurg Psychiatry. 2018;89(12):1308–19.

75. Bhatnagar S, Panguluri SK, Gupta SK, Dahiya S, Lundy RF, Kumar A. Tumor necrosis factor-alpha regulates distinct molecular pathways and gene networks in cultured skeletal muscle cells. PLoS One. 2010;5(10):e13262.

76. Henningsen J, Pedersen BK, Kratchmarova I. Quantitative analysis of the secretion of the MCP family of chemokines by muscle cells. Mol Biosyst. 2011;7(2):311–21.

77. Gomarasca M, Banfi G, Lombardi G. Myokines: The endocrine coupling of skeletal muscle and bone. Adv Clin Chem. 2020;94:155-218.

78. Pedersen BK, Febbraio MA. Muscles, exercise and obesity: skeletal muscle as a secretory organ. Nat Rev Endocrinol. 2012;8(8):457–65.

79. Kieseier BC, Tani M, Mahad D, Oka N, Ho T, Woodroofe N, et al. Chemokines and chemokine receptors in inflammatory demyelinating neuropathies: a central role for IP-10. Brain. 2002;125(Pt 4):823-34.

80. Jubelt B, Cashman NR. Neurological manifestations of the post-polio syndrome. Crit Rev Neurobiol. 1987;3(3):199–220.

81. Salminen A, Kaarniranta K, Kauppinen A. Inflammaging: disturbed interplay between autophagy and inflammasomes. Aging (Albany NY). 2012;4(3):166–75.

82. Iyer SS, Cheng G. Role of interleukin 10 transcriptional regulation in inflammation and autoimmune disease. Crit Rev Immunol. 2012;32(1):23–63.

83. Dorner BG, Scheffold A, Rolph MS, Huser MB, Kaufmann SH, Radbruch A, et al. MIP-1alpha, MIP-1beta, RANTES, and ATAC/lymphotactin function together with IFN-gamma as type 1 cytokines. Proc Natl Acad Sci U S A. 2002;99(9):6181–6.

84. Hardcastle SL, Brenu EW, Johnston S, Nguyen T, Huth T, Ramos S, et al. Serum Immune Proteins in Moderate and Severe Chronic Fatigue Syndrome/Myalgic Encephalomyelitis Patients. Int J Med Sci. 2015;12(10):764–72.

85. Kohno S, Ueji T, Abe T, Nakao R, Hirasaka K, Oarada M, et al. Rantes secreted from macrophages disturbs skeletal muscle regeneration after cardiotoxin injection in Cbl-b-deficient mice. Muscle Nerve. 2011;43(2):223–9.

86. Hornig M, Gottschalk G, Peterson DL, Knox KK, Schultz AF, Eddy ML, et al. Cytokine network analysis of cerebrospinal fluid in myalgic encephalomyelitis/chronic fatigue syndrome. Mol Psychiatry. 2016;21(2):261–9.

87. Haeberle HA, Kuziel WA, Dieterich HJ, Casola A, Gatalica Z, Garofalo RP. Inducible expression of inflammatory chemokines in respiratory syncytial virus-infected mice: role of MIP-1alpha in lung pathology. J Virol. 2001;75(2):878–90.

88. Azzurri A, Sow OY, Amedei A, Bah B, Diallo S, Peri G, et al. IFN-gamma-inducible protein 10 and pentraxin 3 plasma levels are tools for monitoring inflammation and disease activity in Mycobacterium tuberculosis infection. Microbes Infect. 2005;7(1):1–8.

89. Ambrose N, Khan E, Ravindran R, Lightstone L, Abraham S, Botto M, et al. The exaggerated inflammatory response in Behcet’s syndrome: identification of dysfunctional post-transcriptional regulation of the IFN-gamma/CXCL10 IP-10 pathway. Clin Exp Immunol. 2015;181(3):427–33.

90. Reiberger T, Aberle JH, Kundi M, Kohrgruber N, Rieger A, Gangl A, et al. IP-10 correlates with hepatitis C viral load, hepatic inflammation and fibrosis and predicts hepatitis C virus relapse or nonresponse in HIV-HCV coinfection. Antivir Ther. 2008;13(8):969–76.

91. Won EJ, Choi JH, Cho YN, Jin HM, Kee HJ, Park YW, et al. Biomarkers for discrimination between latent tuberculosis infection and active tuberculosis disease. J Infect. 2017;74(3):281–93.

92. Fallahi P, Elia G, Bonatti A. Leishmaniasis and IFN-gamma dependent chemokines. Clin Ter. 2016;167(5):e117-e22.

93. Medoff BD, Sauty A, Tager AM, Maclean JA, Smith RN, Mathew A, et al. IFN-gamma-inducible protein 10 (CXCL10) contributes to airway hyperreactivity and airway inflammation in a mouse model of asthma. J Immunol. 2002;168(10):5278–86.

94. Antonelli A, Ferrari SM, Giuggioli D, Ferrannini E, Ferri C, Fallahi P. Chemokine (C-X-C motif) ligand (CXCL)10 in autoimmune diseases. Autoimmun Rev. 2014;13(3):272–80.

95. Yamagiwa Y, Asano M, Kawasaki Y, Korenaga M, Murata K, Kanto T, et al. Pretreatment serum levels of interferon-gamma-inducible protein-10 are associated with virologic response to telaprevir-based therapy. Cytokine. 2016;88:29-36.

96. Limothai U, Chuaypen N, Khlaiphuengsin A, Posuwan N, Wasitthankasem R, Poovorawan Y, et al. Association of interferon-gamma inducible protein 10 polymorphism with treatment response to pegylated interferon in HBeAg-positive chronic hepatitis B. Antivir Ther. 2016;21(2):97–106.

97. Thanapirom K, Suksawatamnuay S, Sukeepaisarnjaroen W, Tangkijvanich P, Treeprasertsuk S, Thaimai P, et al. Association between CXCL10 and DPP4 Gene Polymorphisms and a Complementary Role for Unfavorable IL28B Genotype in Prediction of Treatment Response in Thai Patients with Chronic Hepatitis C Virus Infection. PLoS One. 2015;10(9):e0137365.

98. Warke RV, Becerra A, Zawadzka A, Schmidt DJ, Martin KJ, Giaya K, et al. Efficient dengue virus (DENV) infection of human muscle satellite cells upregulates type I interferon response genes and differentially modulates MHC I expression on bystander and DENV-infected cells. J Gen Virol. 2008;89(Pt 7):1605-15.

99. Kim J, Choi JY, Park SH, Yang SH, Park JA, Shin K, et al. Therapeutic effect of anti-C-X-C motif chemokine 10 (CXCL10) antibody on C protein-induced myositis mouse. Arthritis Res Ther. 2014;16(3):R126.

100. Misiak B, Bartoli F, Carra G, Malecka M, Samochowiec J, Jarosz K, et al. Chemokine alterations in bipolar disorder: A systematic review and meta-analysis. Brain Behav Immun. 2020.

101. Park H, Poo MM. Neurotrophin regulation of neural circuit development and function. Nat Rev Neurosci. 2013;14(1):7–23.

102. Tang Z, Arjunan P, Lee C, Li Y, Kumar A, Hou X, et al. Survival effect of PDGF-CC rescues neurons from apoptosis in both brain and retina by regulating GSK3beta phosphorylation. J Exp Med. 2010;207(4):867–80.

103. Zembron-Lacny A, Dziubek W, Wolny-Rokicka E, Dabrowska G, Wozniewski M. The Relation of Inflammaging With Skeletal Muscle Properties in Elderly Men. Am J Mens Health. 2019;13(2):1557988319841934.

104. Banerjee C, Ulloor J, Dillon EL, Dahodwala Q, Franklin B, Storer T, et al. Identification of serum biomarkers for aging and anabolic response. Immun Ageing. 2011;8(1):5.

105. Cheng Y, Desse S, Martinez A, Worthen RJ, Jope RS, Beurel E. TNFαlpha disrupts blood brain barrier integrity to maintain prolonged depressive-like behavior in mice. Brain Behav Immun. 2018;69:556-67.

106. Centola M, Cavet G, Shen Y, Ramanujan S, Knowlton N, Swan KA, et al. Development of a multibiomarker disease activity test for rheumatoid arthritis. PLoS One. 2013;8(4):e60635.

